# Development and validation of a framework to improve neglected tropical diseases surveillance and response at the sub-national level in Kenya

**DOI:** 10.1101/2021.05.18.21256594

**Authors:** Arthur K. S. Ng’etich, Kuku Voyi, Clifford M. Mutero

## Abstract

**Background:** Assessment of surveillance and response system functions focusing on notifiable diseases has widely been documented in literature. However, there is limited focus on diseases targeted for elimination or eradication, particularly preventive chemotherapy neglected tropical diseases (PC-NTDs). There are limited strategies to guide strengthening of surveillance and response system functions concerning PC-NTDs. The aim of this study was to develop and validate a framework to improve surveillance and response to PC-NTDs at the sub-national level in Kenya.

**Methods:** Framework development adopted a multi-phased approach. The first phase involved a systematic literature review of surveillance assessment studies conducted in Africa to derive generalised recommendations. The second phase utilised primary data surveys to identify disease-specific recommendations to improve PC-NTDs surveillance in Kenya. The third phase utilised a Delphi survey to assess stakeholders’ consensus on feasible recommendations. The fourth phase drew critical lessons from existing conceptual frameworks. The final validated framework was based on resolutions and inputs from concerned stakeholders.

**Results:** Framework components constituted inputs with the first domain combining surveillance tools, equipment and infrastructure while the second domain combined financial, technical and logistical support. Processes were categorised into four sub-domains with activities for strengthening existing surveillance tools, surveillance core, support and attribute functions. The intended results phase comprised of ten distinct outputs with the anticipated outcomes categorised into three sub domains. Lastly, the overall impact alluded to reduced disease burden, halted disease transmission and reduced costs for implementing treatment interventions to achieve PC-NTDs control and elimination.

**Conclusion:** In view of the mixed methodological approach used to develop the framework coupled with further inputs and consensus among concerned stakeholders, the validated framework appears to be relevant in guiding decisions by policy makers to strengthen the existing surveillance and response system functions towards achieving PC-NTDs elimination.

**Author summary:** Neglected tropical diseases (NTDs) affect marginalised and underserved populations with sub- national levels providing first contact healthcare services to the afflicted communities. NTDs amenable to chemoprophylaxis are primarily controlled through mass treatment interventions. However, identification of disease transmission hotspots requires strengthened health information systems (HIS) to inform targeted public health action and response. Using a multi-phased approach, we developed and validated a framework, which provided a logical approach for guiding actions to strengthen surveillance system functions in view of NTDs. Framework development involved undertaking a systematic literature review to retrieve generalised recommendations for improving surveillance system functions within the African context, conducting primary data surveys to identify disease-specific recommendations on improving surveillance system core, support and attribute functions regarding NTDs and determining feasibility for implementing recommended actions at the sub-national levels. A review of relevant conceptual frameworks provided information underpinning overall framework development. The study identified framework component interlinkages to achieve the desired results of reduced costs for implementing treatment interventions, halted disease transmission and reduced disease burden. Overall, the framework provides a logical approach for strengthening HIS at sub-national levels in NTD endemic regions, considering stakeholders’ perspectives and the available resources to achieve the ultimate goal of disease elimination.

## Introduction

An integral component to achieving health systems strengthening is through enhanced HIS [1, 2]. Quality health information underpins stakeholders’ actions towards achieving quality health care [3]. Essentially, well-functioning health information and surveillance systems ensure generation, analysis, distribution and utilisation of reliable information to support decision-making across all health system levels [4]. A priority action outlined in the Kenyan HIS policy framework is the integration of data collection and dissemination through partnership in health information processes involving all health service providers [2]. This closely relates to the principle of consolidated efforts within the integrated disease surveillance and response (IDSR) system framework adopted by World Health Organization (WHO) Member States in Africa [5]. The IDSR framework categorises diseases as either being epidemic-prone, diseases of public health importance or diseases targeted for elimination or eradication [5]. In particular, neglected tropical diseases (NTDs) in Kenya are categorised as either being of public health importance or targeted for elimination or eradication [6]. NTDs are a diverse group of communicable diseases that prevail in tropical and subtropical conditions, mainly affecting the marginalised communities and are a major cause of deformities and disabilities [7]. The WHO work plan for NTDs elimination identifies five intervention strategies including preventive chemotherapy among affected populations, intensified case finding and disease management, integrated vector management, improved sanitation and hygiene and veterinary public health [8]. Preventive chemotherapy (PC) is an essential strategy for NTDs infection control, which targets the disease causative agents – parasitic, viral and bacterial – concurrently through mass drug administration (MDA) [9]. However, focus on MDA strategies limits priority given to the role played by other social determinants and the existing health systems to achieve NTDs control. There is consensus among NTD experts that in order to achieve efficient and sustainable control and meet disease-specific elimination targets it requires well-functioning health systems [10–13]. Overdependence on mass treatment strategies may exhaust the available resources, thereby impeding health system strengthening efforts [10–12].

Neglected tropical diseases are a clear case in point regarding existing health systems functioning sub-optimally. In 2013, the sixty-sixth World Health Assembly adopted a resolution on strengthening disease surveillance systems specific to NTDs targeted for eradication and supporting Member States in their efforts to manage data generated within national surveillance systems [14]. NTDs are considered of low priority by national HIS and within disease surveillance systems, which may hinder generation of quality data and result to ineffective use and dissemination of information [14]. This identifies the need to establish strong and adaptable HIS to improve data management capacities in NTD endemic countries [14]. Strengthened HIS will facilitate undertaking evidence-based actions and establishment of effective NTDs control interventions [10, 11, 14]. Existing interventions and technologies may not solely achieve sustainable control and elimination of targeted NTDs [11, 15]. Therefore, identifying the need to strengthen the current surveillance and response systems to inform effective and targeted control strategies in endemic regions. Furthermore, since most preventive chemotherapy neglected tropical diseases (PC-NTDs) are endemic in resource constrained countries, enormous gaps in the area of surveillance-response systems in such settings can only be overcome through renewed efforts of applied research. In addition, this could be complemented by strengthening existing health systems as well as developing effective and novel frameworks tailored to local settings [16]. Nevertheless, strengthened surveillance system functions specific to PC-NTDs would have a positive snowball effect towards response actions for other conditions considering an integrated disease surveillance and response approach.

The current framework was partly founded on principles adopted by conceptual frameworks for strengthening public health surveillance systems [17, 18]. The intricacies of strengthening health system building blocks are evident from the varying perspectives illustrated by an array of conceptual frameworks applicable to different settings [19–21]. Additionally, the present framework was context-specific and relied on findings and recommendations emanating from a previous systematic review of literature and studies involving primary data surveys to assess disease surveillance and response activities. Systematic literature reviews provide a valid approach to assessing the feasibility of implementing specific public health practices [22]. On the other hand, results from studies involving primary users of a given HIS are vital for strengthening specific system components. Frameworks developed with the intention of assessing the strength of a health system should typically address all essential components including the resources dedicated to the system (inputs), what the system inputs intend to fulfill (processes and outputs) and the resulting benefits and fundamental change to the system (outcomes and impact) [19]. However, multiple complexities are involved since the overall health system performance is dependent on several other factors involving technical, social, organizational and cultural dynamics [19, 21]. The framework herein focuses on strengthening the indicator-based components of the existing surveillance system that involves the collection of structured data through routine surveillance systems [23]. The components of the framework are specific to improving surveillance and response to NTDs amenable to chemoprophylaxis.

The chronic nature of NTDs leads to a sustained cycle of poverty among the afflicted communities and exerts pressure on the already fragile health systems [24]. Health system strengthening efforts in NTD endemic regions are paramount to improve health workers’ attitudes and health system trustworthiness among the affected communities [25]. Previously, public health systems performance assessments have prioritised execution of interventions, resulting outcomes and their impact. However, there is limited focus on linking the outcomes to public health system inputs and processes. Therefore, the proposed framework intended to conceptualise the link between inputs, processes and relevant outputs and outcomes of the existing surveillance system in relation to PC-NTDs. The framework was based on stakeholders’ perspectives on improving surveillance and response to PC-NTDs, with the intention of strengthening implementation of surveillance activities at the county level in Kenya. The goal was to enable concerned stakeholders effectively identify gaps within the existing surveillance system, prioritise plans of actions for improving the system and design indicators for progress monitoring [26]. The framework is intentional to facilitate planning of PC-NTDs surveillance activities with the end-goals in mind to achieve the overall desired outcomes rather than considering inputs and processes independently. Dependence on chemoprophylaxis as a stand-alone intervention for PC-NTDs requires continued support of HIS components through strengthened surveillance systems. Therefore, the proposed framework aims to provide a clear, acceptable and adoptable logical approach to guide decisions and policies concerning surveillance and response to PC-NTDs at the sub national level in Kenya.

## Methods

### Ethics statement

Written informed consent was obtained from research participants involved at various stages of framework development and validation. Ethical approval was granted by the Faculty of Health Sciences Research Ethics Committee of the University of Pretoria in South Africa (**Ethics Reference No: 27/2018**) and the Institutional Research and Ethics Committee (IREC) of Moi University/Moi Teaching and Referral Hospital in Kenya (**Formal Approval No: IREC 2099**). In addition, the National Commission for Science, Technology and Innovation (NACOSTI) provided research authorisation to undertake the research in Kenya (**Reference No: NACOSTI/P/18/62894/21393**). Permission to conduct the overall study was granted by the Ministry of Health in Kenya and relevant County health authorities.

### Methodology for framework development

Development of the framework for improving PC-NTDs surveillance and response encompassed four main phases (Fig 1). The first phase involved conducting a systematic review of literature, which identified generalised but critical recommendations emerging from prior studies to improve disease surveillance and response systems in the African region. The second phase involved primary data surveys conducted in PC-NTD endemic regions in Kenya to assess performance of surveillance core, support and attribute functions. The third phase used a modified-Delphi survey to assess stakeholders’ consensus on the importance and feasibility of implementing recommended actions to improve PC-NTD surveillance and response at the sub-national level in Kenya. Lastly, the fourth phase involved a review of previous HIS and public health surveillance system conceptual frameworks to gain insights into the existing frameworks.

**Figure 1.**
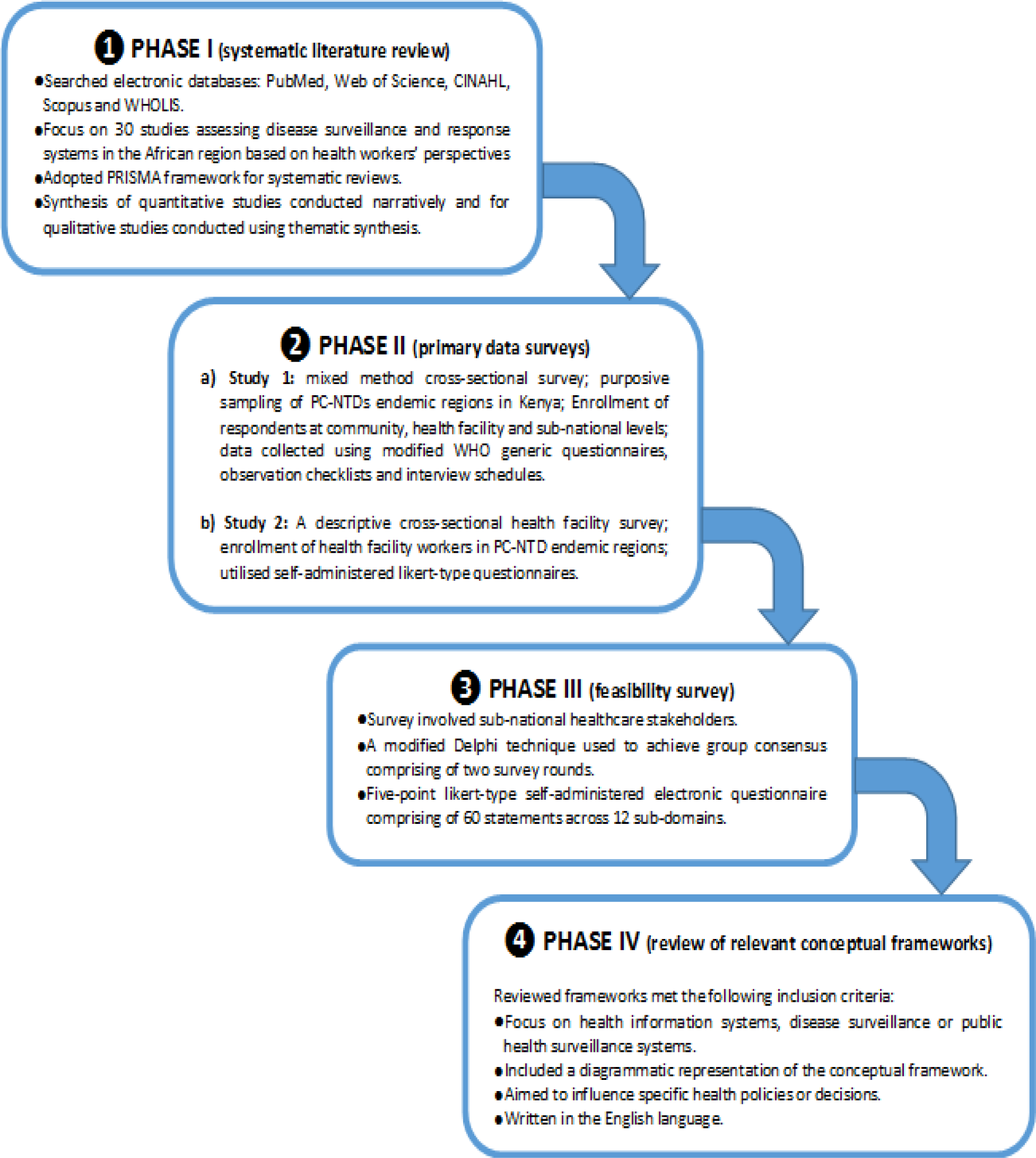
Framework development process.

### Phase I: Systematic review of literature on surveillance system assessment studies [27]

The purpose for this phase was to systematically review both published and grey literature retrieved from the relevant online databases including PubMed, Web of Science, Scopus, Cumulative Index for Nursing and Allied Health Literature (CINAHL) and World Health Organisation Library and Information Networks for Knowledge (WHOLIS). The focus of the systematic literature review process was to search for studies assessing disease surveillance and response systems in the African region based on health workers’ perspectives (PROSPERO Registration number CRD42019124108) [27]. The review focused on the post-adoption phase of the revised IDSR system as from 2010 and onwards. A combination of keywords were used to search for relevant studies including “surveillance”, “public health surveillance”, “integrated disease surveillance and response”, “evaluation”, “assessment”, “health worker”, “healthcare personnel”, “Africa” and “Sub Saharan Africa”. The review aimed to assess key findings and recommendations to improve surveillance and response systems in the African region based on health workers’ perspectives.

### Phase II: Primary data surveys on surveillance core, support and attribute functions concerning PC-NTDs [28, 29]

The aim of this phase was to conduct two empirical studies in NTDs endemic regions in Kenya. The studies aimed to assess performance of surveillance functions concerning PC-NTDs as perceived by healthcare workers. The two studies intended to put focus on improving PC-NTDs surveillance within the existing IDSR system, which is in line with the strategic objectives outlined in the Second National Strategic Plan for Neglected Tropical Diseases and the National Breaking Transmission Strategy in Kenya [30, 31]. The first study used a mixed method cross-sectional survey to assess surveillance system core and support functions relating to PC-NTDs [28]. The study sites were purposively sampled from three regions in Kenya, which are co-endemic of at least three or more fully mapped PC-NTDs. Health personnel involved in the study were enrolled from the community (50), health facility (192) and sub-national (44) levels. Modified WHO generic questionnaires, observation checklists and interview schedules were used for data collection. The second study aimed to evaluate surveillance system attributes based on health facility workers’ perceptions concerning PC-NTDs [29]. The study sites were similar to those selected in the first survey. The study adopted a descriptive cross-sectional health facility survey involving 192 health workers responsible for disease surveillance and response activities in NTD endemic regions in Kenya. Self-administered likert-type questionnaires were used to assess healthcare personnel perceptions of surveillance system attributes on simplicity, acceptability, stability, flexibility, usefulness, data quality and reporting timeliness and completeness with regard to PC-NTDs.

### Phase III: Feasibility of implementing recommendations to improve PC-NTDs surveillance and response [32]

This third phase was based on a Delphi survey, which involved enrollment of key stakeholders at the sub-national level responsible for overseeing disease surveillance and response activities in Kenya [32]. The survey aimed to assess the principal and feasible recommendations derived from the preceding first and second phase. This phase guided selection of key constructs making up the framework in line with stakeholders’ opinions and inputs. Stakeholders constituted individuals that regularly utilise information generated by the surveillance system to take appropriate actions or make decisions influencing implementation of interventions and policy-making processes [33, 34].

### Phase IV: Review of relevant conceptual frameworks

In the fourth phase, a comprehensive review of existing HIS and public health surveillance system conceptual frameworks was undertaken. Retrieval of the relevant documents involved basic search strategies; electronic database search, a manual reference search of selected documents and use of a generic internet search engine. Literature searches were undertaken using PubMed, Web of Science, Scopus, Google Scholar and a free Google search to identify the relevant frameworks. The key search terms included: “conceptual framework”, “model”, “health systems”, “health information systems”, “public health surveillance systems”, “surveillance and response”. These terms were used in various combinations to ensure the literature search was extensive. A manual reference check of the included frameworks was conducted to identify additional frameworks that met the inclusion criteria. The inclusion criteria for the reviewed frameworks was a specific focus on HIS, disease surveillance or public health surveillance systems; included a diagrammatic representation of the conceptual framework; aimed to influence specific health policies or decisions; and written in the English language. This review involved critical assessment of components making up the frameworks. In addition, reviewing the framework components provided an understanding on the relevant interlinkages, which formed the basis for the proposed framework.

## Results

### Phase I: Systematic review of literature on surveillance system assessment studies [27]

The first phase involved conducting a systematic review of literature of previous surveillance assessment studies in the African region. Thirty studies met the inclusion criteria and were assessed to retrieve generalised recommendations to improve surveillance systems within the revitalised IDSR framework adopted by African countries. Eighteen emerging sub-themes were identified for recommendations specific to improving four core surveillance functions and three support functions. However, no specific sub-themes emerged from assessing the surveillance system attributes. The surveillance core functions included; case confirmation, reporting, data analysis and feedback. Surveillance support functions included; training, supervision and resources while surveillance system attributes included; reporting timeliness and completeness, data quality and accuracy, usefulness, acceptability, simplicity and stability. Other key recommendations alluded to adoption of alternative surveillance strategies and calls for further research to strengthen surveillance systems.

Main themes were based on elements constituting the broader surveillance functions – core, support and attributes – while 18 emerging sub-themes were derived from the recommendations. Surveillance functions themes and sub themes (provided in parentheses) included; case confirmation (improved specimen handling [35] and strengthened laboratory support [36, 37]); reporting (improved reporting quality [35, 38–40] and adequacy of reporting forms provision [35, 41, 42]); feedback (improved health workers’ attitudes [43] and enhanced feedback from higher to lower surveillance levels [44–46]); data analysis (surveillance system performance monitoring [47] and improved data accuracy [46, 48]). On the other hand, key recommendations regarding surveillance support functions included; training (improved surveillance system performance [37, 49, 50], improved surveillance data quality [35, 42, 51–53] and enhanced knowledge on surveillance systems [40, 41, 54–61]); supervision (strengthened implementation of surveillance system activities [41, 47, 50, 60], utilisation of up-to-date information [37] and identification of correct reporting channels [41, 57]); resources (financial resources [39, 41, 47, 50, 52, 60], human resources [37, 59], technical, material and logistical resources [36, 39, 45, 46, 54, 57, 59] and equipment and infrastructure [37, 51, 53, 57]). Recommendations regarding surveillance system attributes were specific to reporting timeliness and completeness [37, 41, 43, 46, 48, 49, 55, 56, 58, 60], data quality and accuracy [48, 49, 55, 56], usefulness [43, 55], acceptability [49, 55, 57], stability [49, 53, 55, 57, 62] and simplicity [43, 49, 55–57]. Other recommendations included; alternative surveillance strategies (electronic based surveillance [37, 43, 49, 56, 57, 63], community based surveillance [47, 58] and syndromic surveillance [63]) and further calls for research on surveillance [48, 49, 59].

### Phase II: Primary data surveys on surveillance core, support and attribute functions concerning PC-NTDs [28, 29]

The first phase described surveillance system assessment studies previously conducted in the African region, which identified broad recommendations to improve the existing surveillance systems; however, these recommendations were inclined to notifiable diseases considered of national priority in African countries. Therefore, based on the premise that diseases targeted for control or elimination were to be accorded priority commensurate to notifiable diseases, the second phase entailed conducting health worker surveys involving primary data collection to assess performance of surveillance core, support and attribute functions concerning PC-NTDs endemic in Kenya.

### Phase II (a) – Assessment of surveillance core and support functions regarding neglected tropical diseases in Kenya [28]

The first study aimed to assess surveillance core and support functions regarding PC-NTDs endemic in Kenya. This identified specific recommendations to strengthen surveillance functions focusing on PC-NTDs within the existing IDSR system. There were eleven a priori identified main themes with up to 62 emerging sub-themes based on recommendations to improve PC-NTDs surveillance and response according to health workers’ perspectives. A high degree of groundedness was defined as sub-themes (recommendations) that were mentioned fifteen or more times (G ≥ 15) by the research participants under each main theme. Recommendations to improve PC-NTDs surveillance core activities were categorised into 7 main themes and 32 sub-themes. The principal recommendations considering a high degree of groundedness included provision of simplified case definitions, improved laboratory capacity, provision of PC-NTDs reporting tools, timely and regular feedback on surveillance reports, adoption of electronic feedback mechanisms, improved data analysis skills and provision of adequate outbreak response supplies. On the other hand, recommendations to strengthen surveillance support activities comprised of 4 main themes and 30 sub-themes. Therefore, the main recommendations based on a high degree of groundedness encompassed provision of surveillance guidelines for supervision, properly constituted supervisory teams, regular supervision from higher levels, frequent provision of PC-NTDs updates, regular sensitisation and involvement of all health workers in surveillance activities, enhanced training on PC-NTDs surveillance, improved human resource capacity, provision of surveillance reporting and analysis tools, availing equipment and training materials, provision of adequate funding to facilitate surveillance activities and provision of reliable transportation.

### Phase II (b) – Evaluation of health surveillance system attributes regarding neglected tropical diseases in Kenya [29]

The subsequent study aimed to evaluate surveillance system attributes based on health workers’ perceptions concerning PC-NTDs endemic in Kenya. Health workers perceived the surveillance system to be simple (55%), acceptable (50%), stable (41%), flexible (41%), useful (51%) and to provide quality surveillance data (25%). Health personnel experienced difficulties completing IDSR reporting forms with regard to PC-NTDs, easily understanding reporting guidelines and utilising PC-NTDs case definitions. Further, health workers were less satisfied with their involvement in facility-based surveillance activities. Health workers perceived PC-NTDs to be of low priority compared to other conditions and were only willing to be involved in surveillance activities of diseases considered of priority. However, community level health workers were more willing to support and be involved in PC-NTDs surveillance activities. Heath workers felt the existing IDSR system lacked flexibility with regard to being well adapted to report all PC-NTDs co-endemic in the region and easily adapting to changes in information needs, technological shift and funding. Furthermore, minimal adaptability of the existing surveillance system to changes in reporting mechanisms from paper-based to electronic systems was reported. Respondents further indicated low stability of the surveillance system in terms of adequacy of the available forms to report PC-NTDs, resource sufficiency and capacity of health managers to support surveillance activities and address challenges promptly. Health workers also reported that the surveillance system lacked usefulness in terms of generating sufficient PC-NTDs surveillance data to inform implementation of control interventions and influence sufficient donor support. In addition, respondents felt the surveillance system lacked capacity to provide adequate information to inform efficient public health actions in view of PC-NTDs. Health facilities sampled from the PC-NTDs endemic regions hardly met the 80% target reporting thresholds. Only about one-third of the facilities met this threshold in terms of reporting timeliness of monthly surveillance data and slightly more than a half of facilities met the threshold for completeness of monthly reports. Overall, considering a cut off attribute score of above 50%, findings showed health workers perceived the existing surveillance system to be simple, acceptable and useful with regard to PC-NTDs. However, respondents had low perceptions on the stability, flexibility and data quality of the surveillance system. Notably, there was a dwindling trend in monthly reporting timeliness and completeness rates by health facilities over a three-year period.

### Phase III: Feasibility of implementing recommendations to improve PC-NTDs surveillance and response [32]

The first and second phase provided a plethora of recommendations on improving the existing surveillance systems considering broad perspectives within the African context and further actions peculiar to improving PC-NTDs surveillance and response within an endemic setting [27–29]. Consequently, the third phase integrated evidence from the systematic literature review with findings from the primary data surveys to generate actionable statements that were assessed by relevant health stakeholders on their importance and feasibility for implementation at the sub- national level in Kenya. Findings from the Delphi survey indicated that stakeholders agreed on the importance of 56 recommendation statements and further reached consensus on the feasibility of implementing 47 recommended actions at the sub-national level. Stakeholders had a converging opinion on the importance and feasibility of implementing recommendations in six sub-domains relating to feedback, epidemic preparedness and response and those relating to four surveillance system attributes – simplicity, acceptability, stability and flexibility. However, there was lack of consensus on specific recommendations regarding the remaining six sub-domains on their importance or feasibility for implementation. Furthermore, sensitivity analysis that incorporated those participants with neutral responses indicated that the participants considered all recommendation statements to be of importance. Nevertheless, there was still non-consensus on the feasibility of availing disease-specific case registers, confirmation of all PC-NTDs cases, undertaking routine data analysis, increasing the number of supervisory visits at the lower levels, involving all health workers in surveillance training, retaining trained surveillance personnel and increasing the number of health workers responsible for overseeing surveillance activities at the sub-national level.

### Phase IV: Review of relevant conceptual frameworks

The systematic search identified 10 records eligible for full-text review. However, only 6 distinct conceptual frameworks focusing on HIS and public health surveillance systems met the inclusion criteria for review. Critical lessons drawn from HIS conceptual frameworks alluded to stakeholders’ involvement, the organisational setting and technological aspects. Further, reviewed public health surveillance frameworks focused on performance indicators, disease-specific surveillance aspects and outbreak detection. The proposed framework was founded upon the Health Metrics Network (HMN) framework, which recognises the critical role of stakeholders to reach consensus on decisions to strengthen HIS [19]. A pertinent component of the HMN framework focuses on inputs, processes, outputs and outcomes of HIS. These specifically includes resources, indicators, data sources, data management, dissemination and use of information by stakeholders to guide the decision-making process [19]. The framework further identifies that evaluation of HIS can be challenging considering other technical, social, organisational and cultural aspects [19]. Furthermore, the Performance of Routine Information System Management (PRISM) framework assesses HIS performance considering technical, organisational and behavioral factors [20]. The PRISM framework guided development of the current framework since it identifies inputs based on the technical, behavioral and organizational elements, which determine data management (processes) and consequentially influence data quality and utilisation (outputs), health system performance (outcomes), and net impact on health status [20]. The proposed framework further drew on the concept of “fit” based on the human, organisation and technology fit model (HOT-fit), which is concerned with HIS ability, stakeholders’ practices and the organisational setting aligned with each other [21]. The framework describes various interdependent dimensions considering how system, information and service quality work independently or in concert with one another to influence system use and users’ satisfaction. Moreover, the framework illustrates how the organisational structure and environment work jointly to impact system use and overall net benefits [21].

The current framework further adopted core and support activities indicators outlined in the conceptual framework authored by McNabb et al. [17]. The framework links surveillance activities to the resulting public health actions [17]. This framework intended to aid healthcare practitioners to design future models through a series of activities to achieve measurable outputs and outcomes [17]. Furthermore, the authors posited that the framework would be practical in developing countries undergoing both political and economic reforms having disease surveillance systems that function sub-optimally. Similarly, the proposed framework adopted comparable concepts described in a disease-specific framework informed by stakeholders’ inputs to identify either process-oriented or outcome-oriented performance indicators for evaluating surveillance core and support activities [18]. Additionally, the current framework aimed to improve epidemic preparedness and response actions, which were relatable to the processes and outputs contained in a framework for evaluating public health surveillance systems for early detection of outbreaks [34]. Lessons learned from the reviewed conceptual frameworks alluded to stakeholders’ involvement, framework components and their interlinkage and disease-focused perspectives.

### Development of a framework to improve PC-NTDs surveillance and response

Framework development was informed by feasible recommendations derived from the third phase; in addition, to the lessons drawn from reviewing relevant conceptual frameworks. The proposed framework aimed to provide a logical approach hinged on the inputs, processes and outputs to enable decision makers institute evidence-based actions to achieve the intended outcomes and the desired impact [64]. A stakeholder-oriented approach was adopted throughout the framework development process to identify practically feasible recommendations for implementation and ensure utilisation of evidence-based research findings for decision-making [65]. The underlying assumption was that research findings can only be reliable to effectively influence the policy making process if they are endorsed by concerned stakeholders [65]. This assumption formed the basis for undertaking the third phase, which identified key components of the proposed framework. We also adopted the logical framework approach (LFA) as stipulated in the guide to monitoring and evaluating communicable disease surveillance and response systems [66]. The framework components were categorised based on their focus on input factors (resources), process factors (planned activities), output factors (short, medium and long-term benefits), outcome factors (intended results) and the overall impact factors [66]. Framework development was guided by findings from the first, second, third and fourth phases with the aid of a logical component matrix to identify the interrelation between the derived themes (Table 1). The proposed framework intended to improve the existing surveillance and response system focusing on PC-NTDs, which will in turn strengthen implementation of the core medical interventions for PC-NTDs control; preventive chemotherapy and intensified case finding and disease management [8, 67].

**Table 1.**
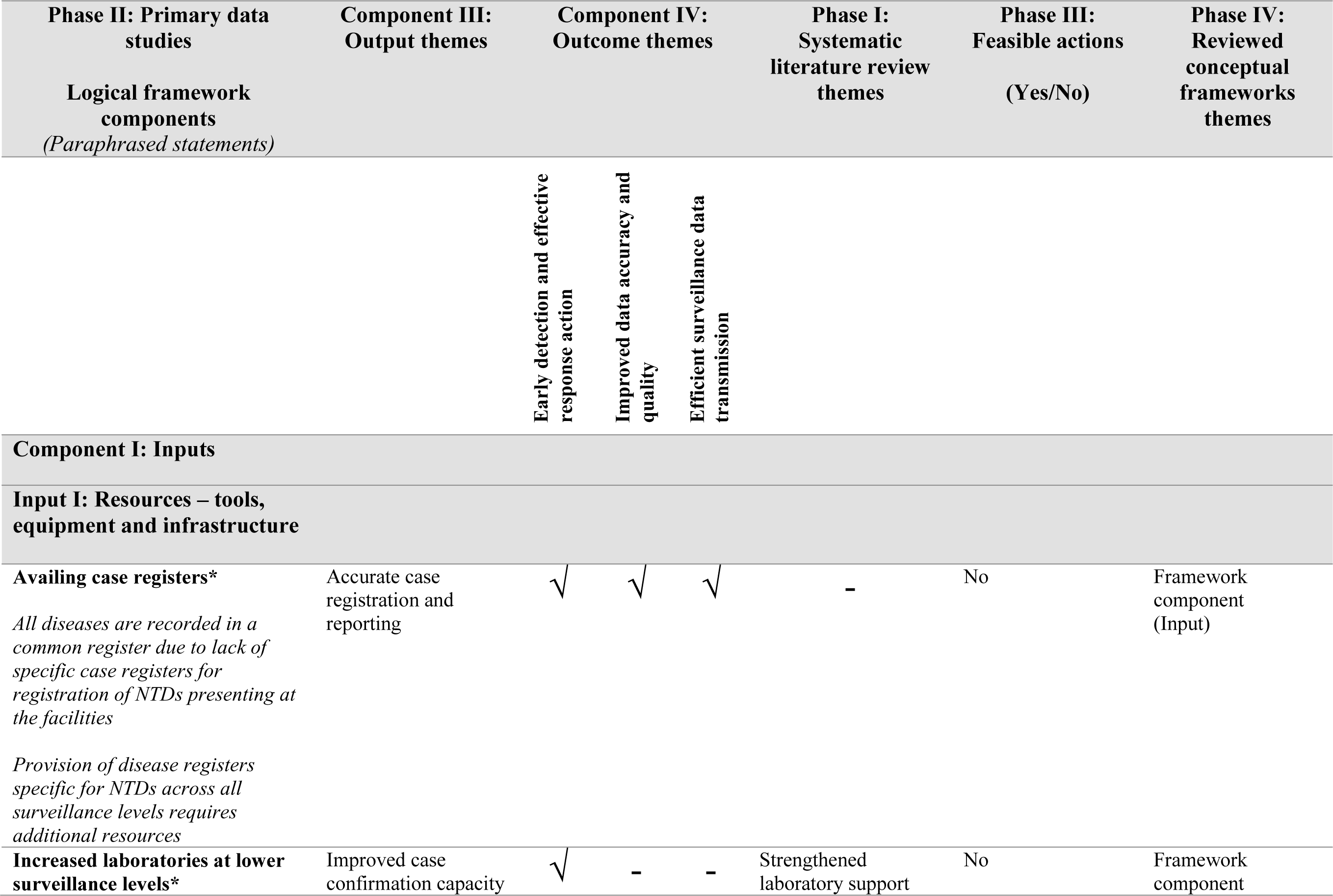
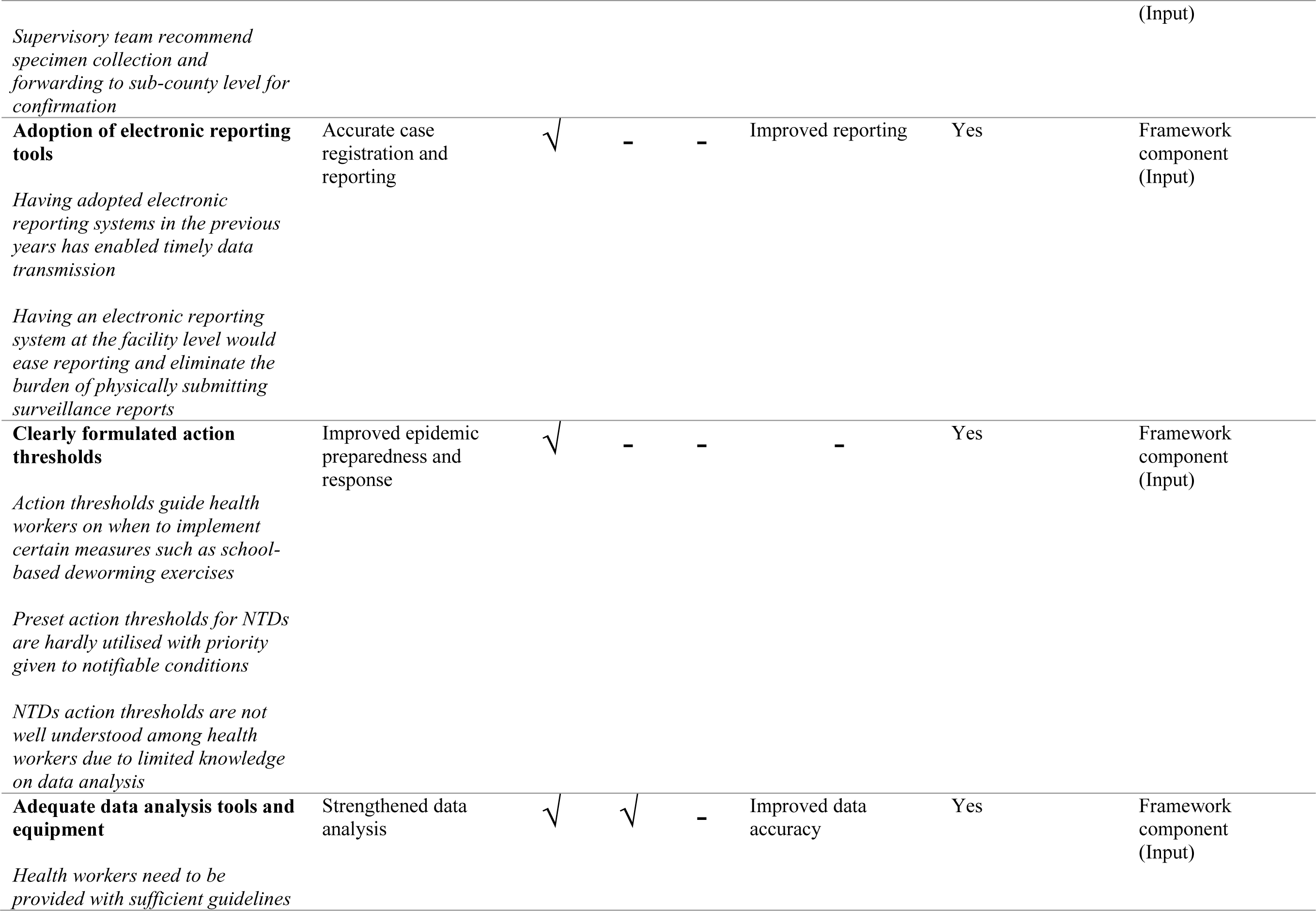
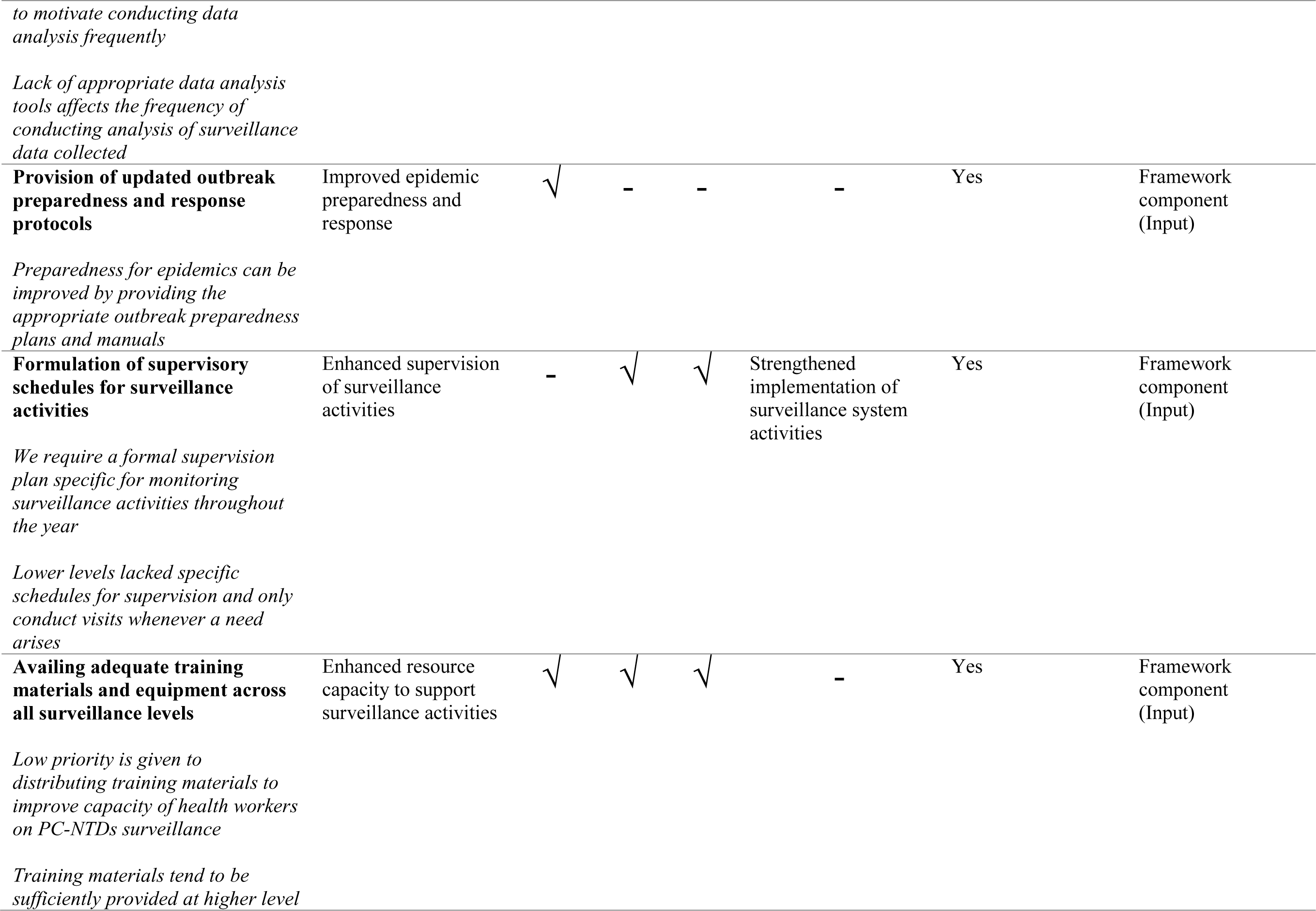
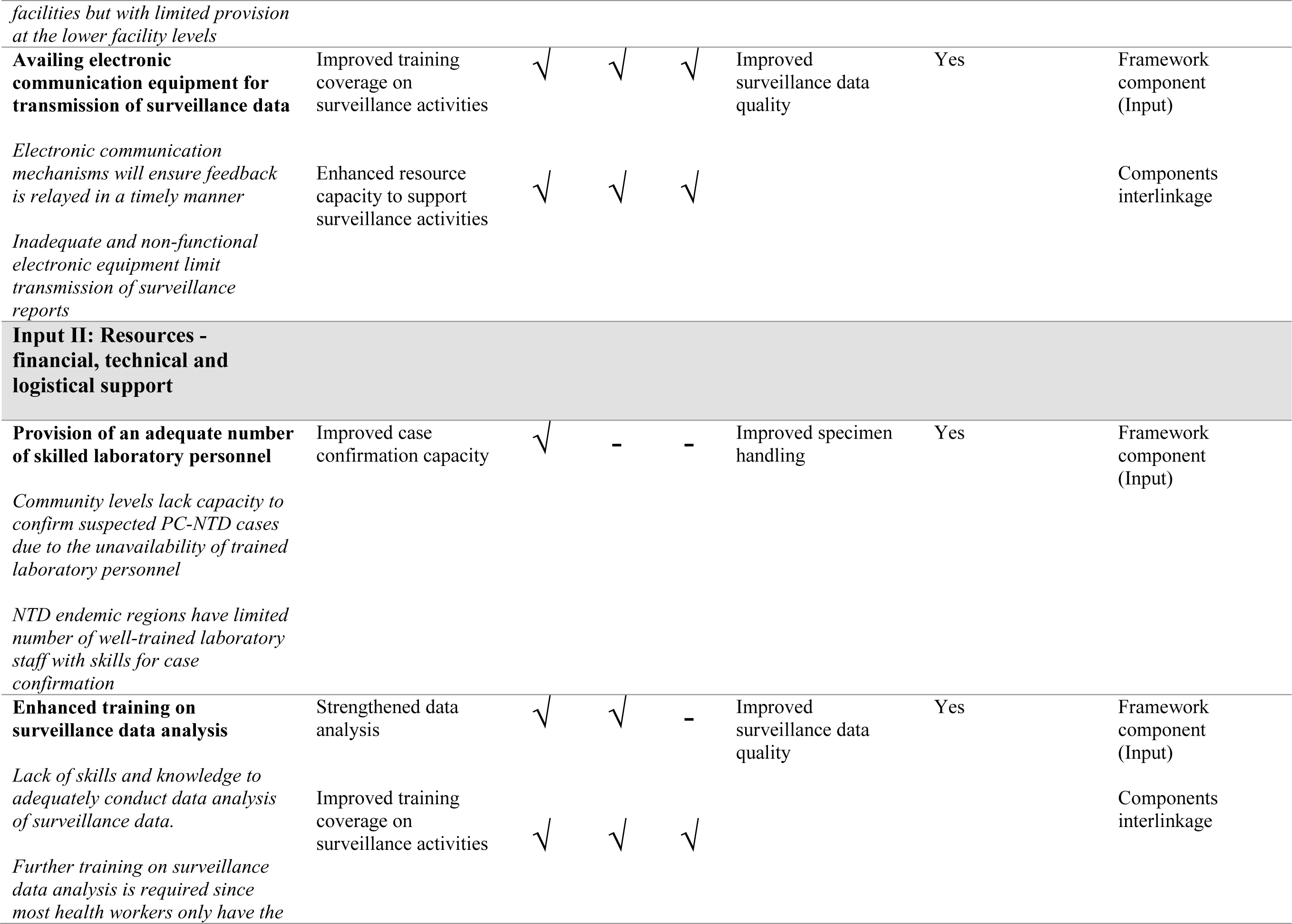
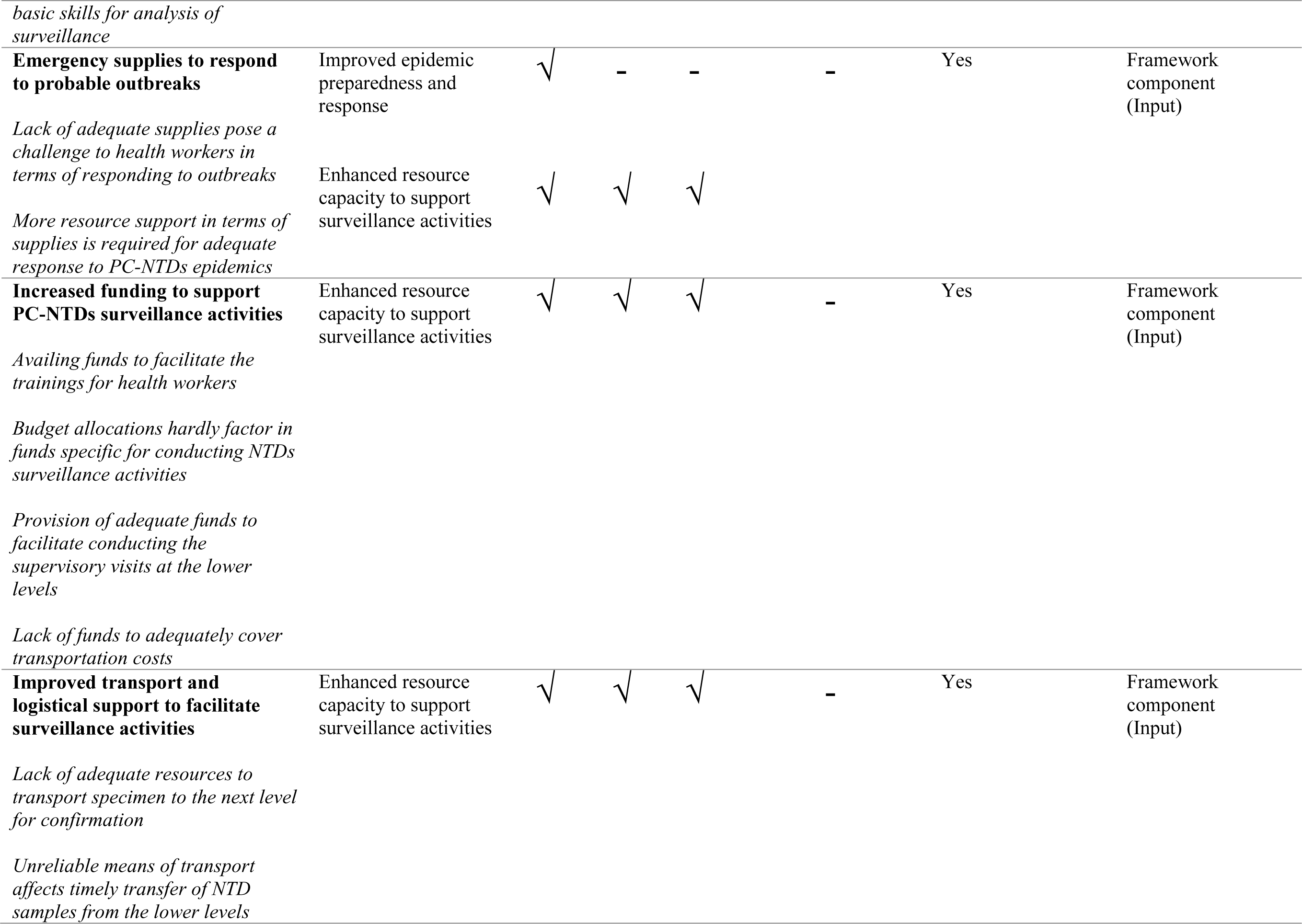
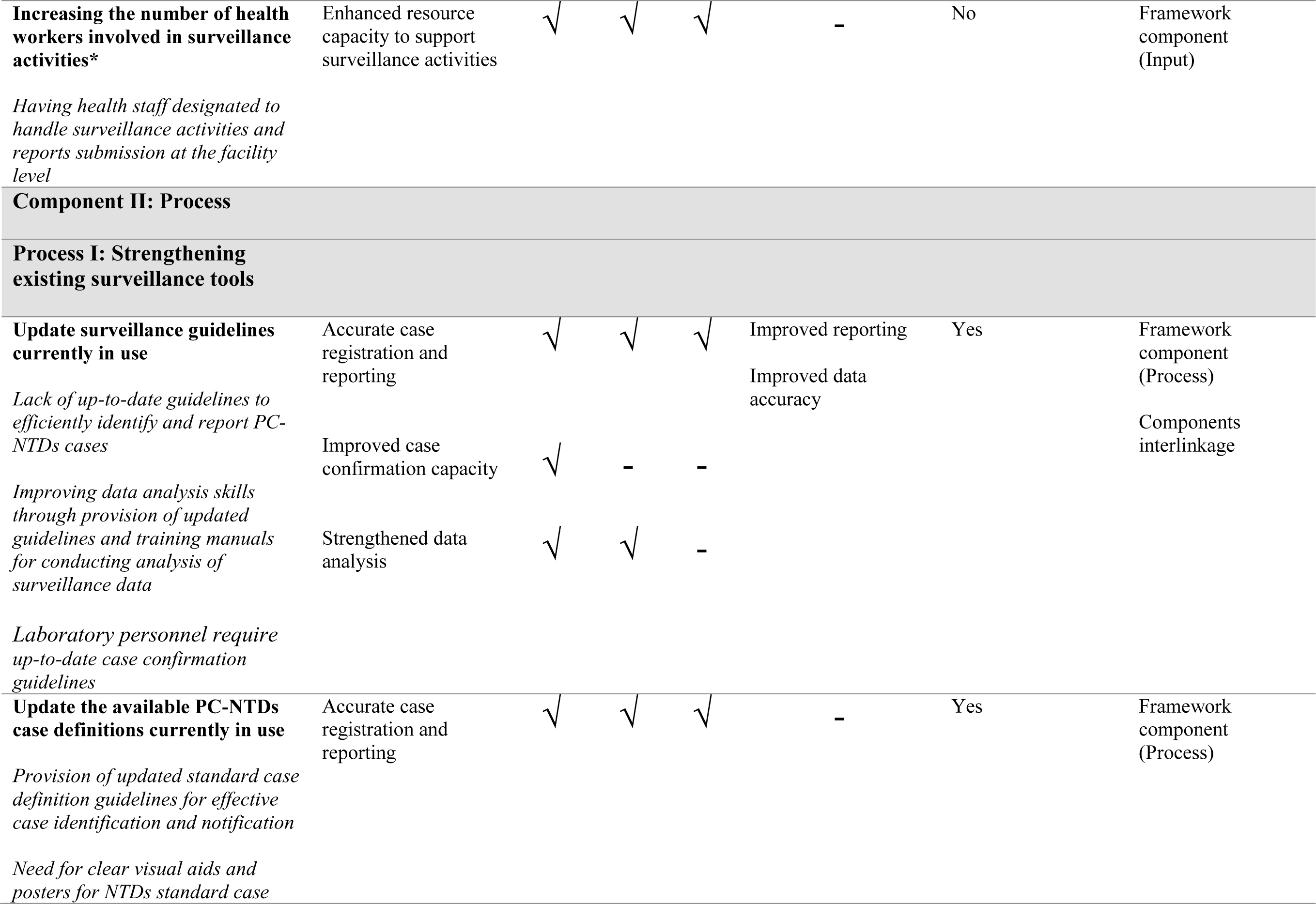
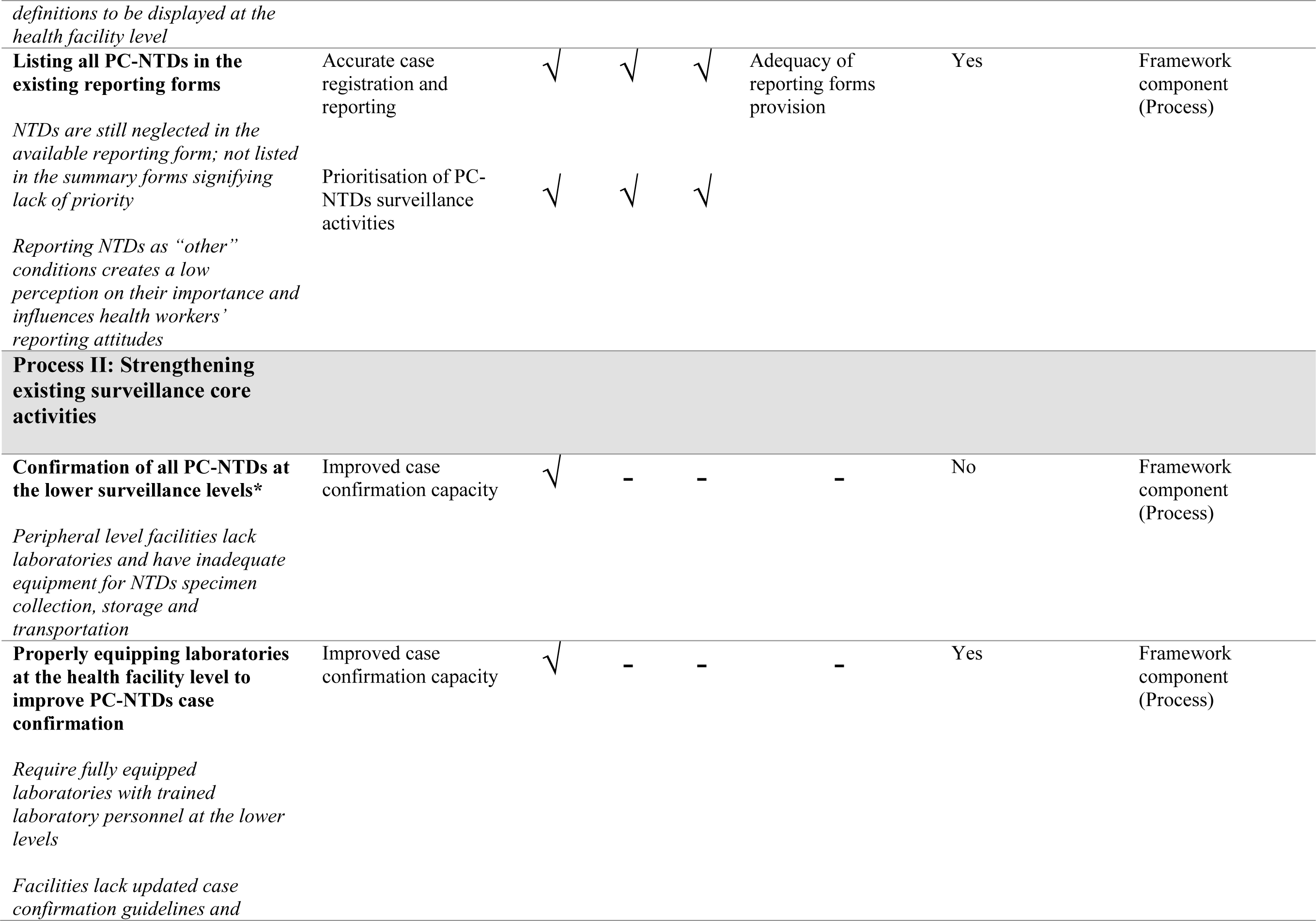
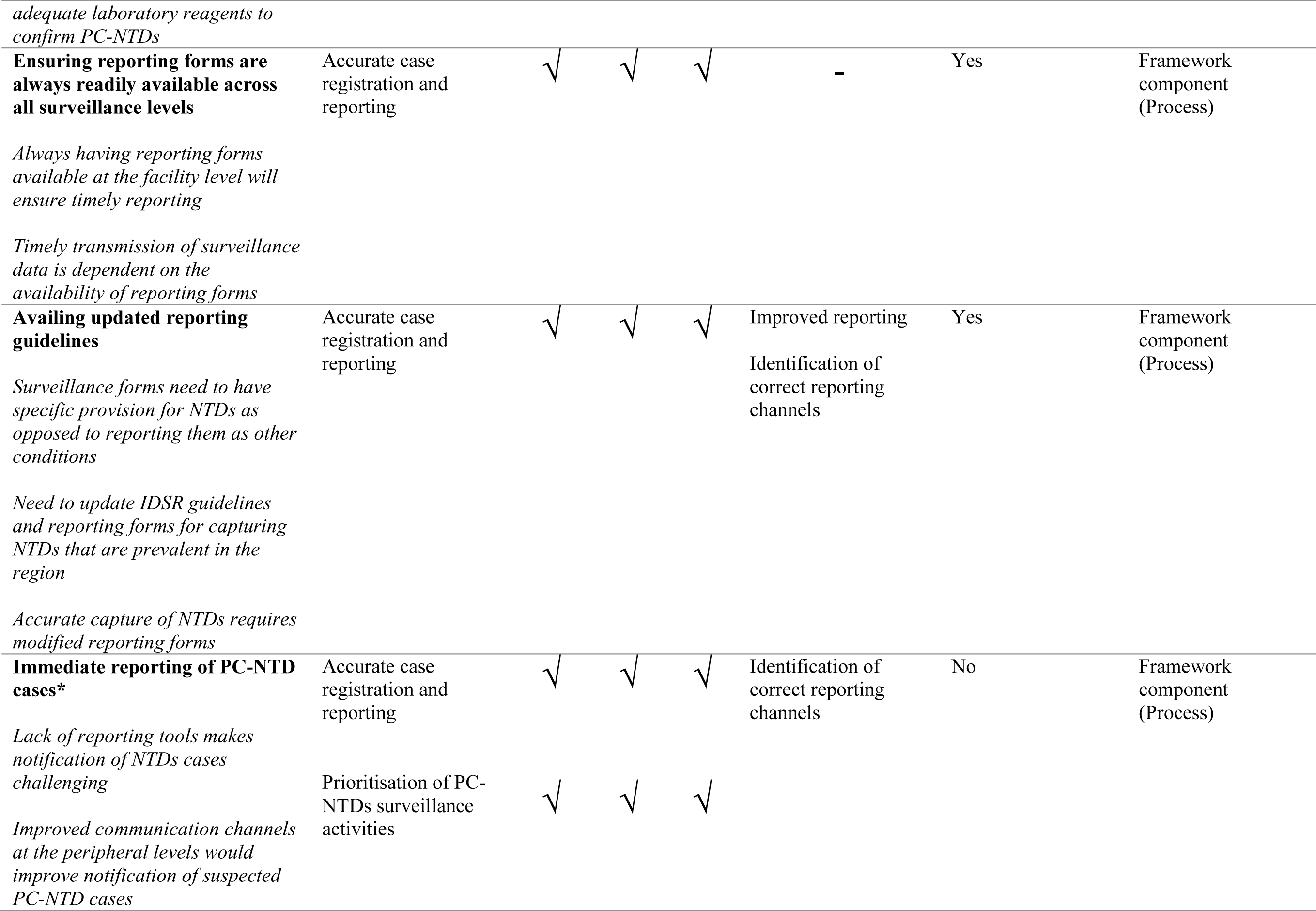
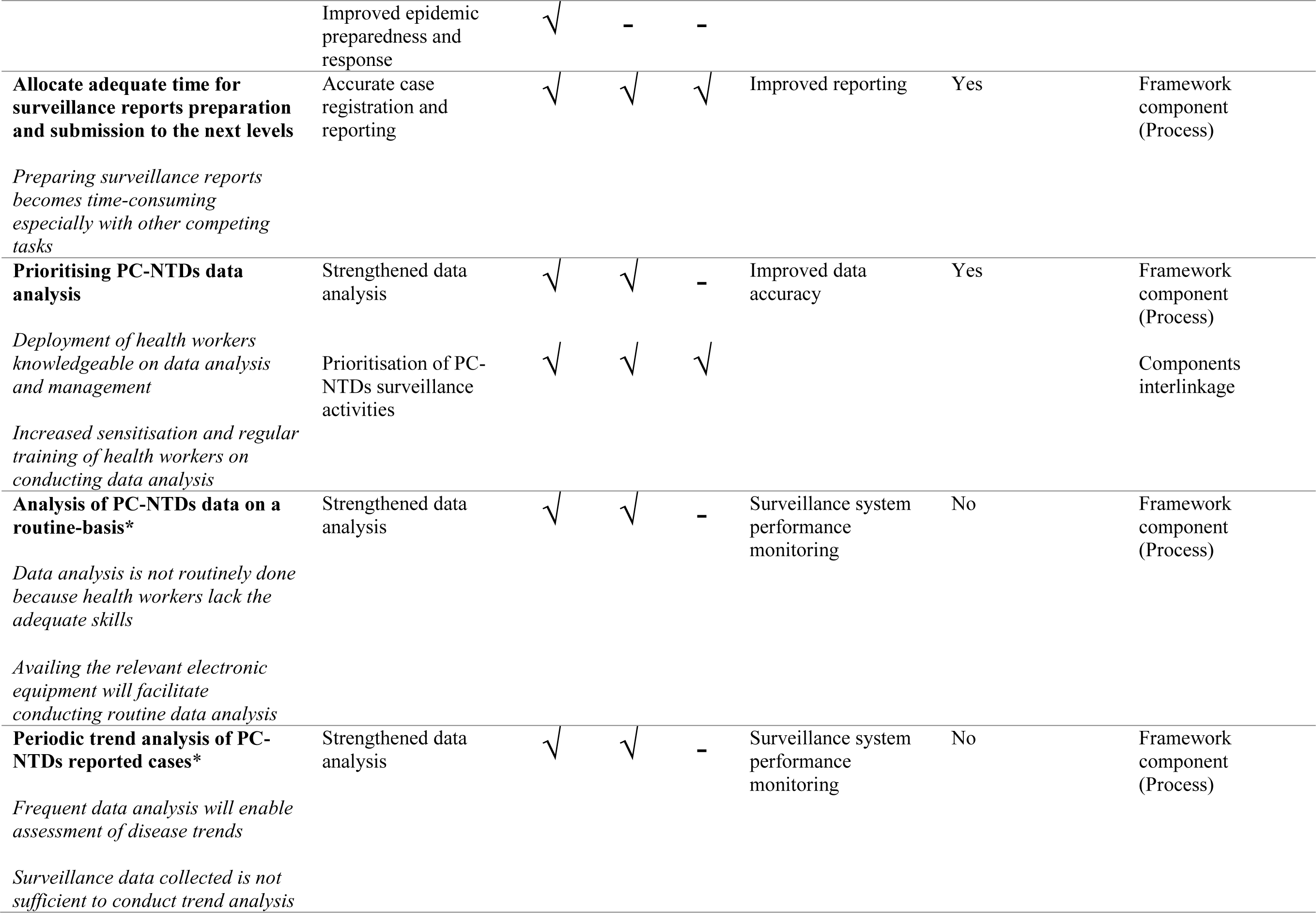
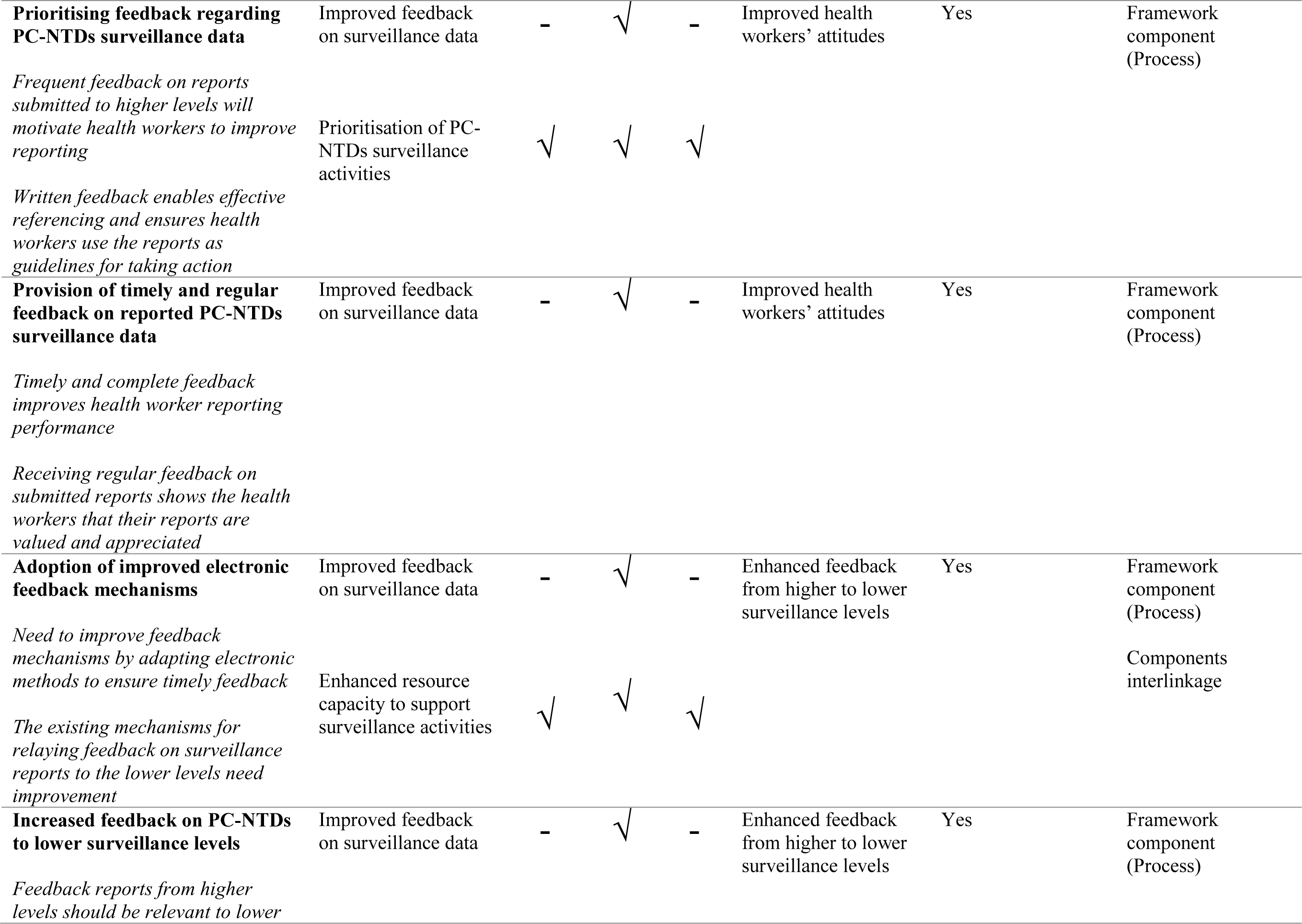
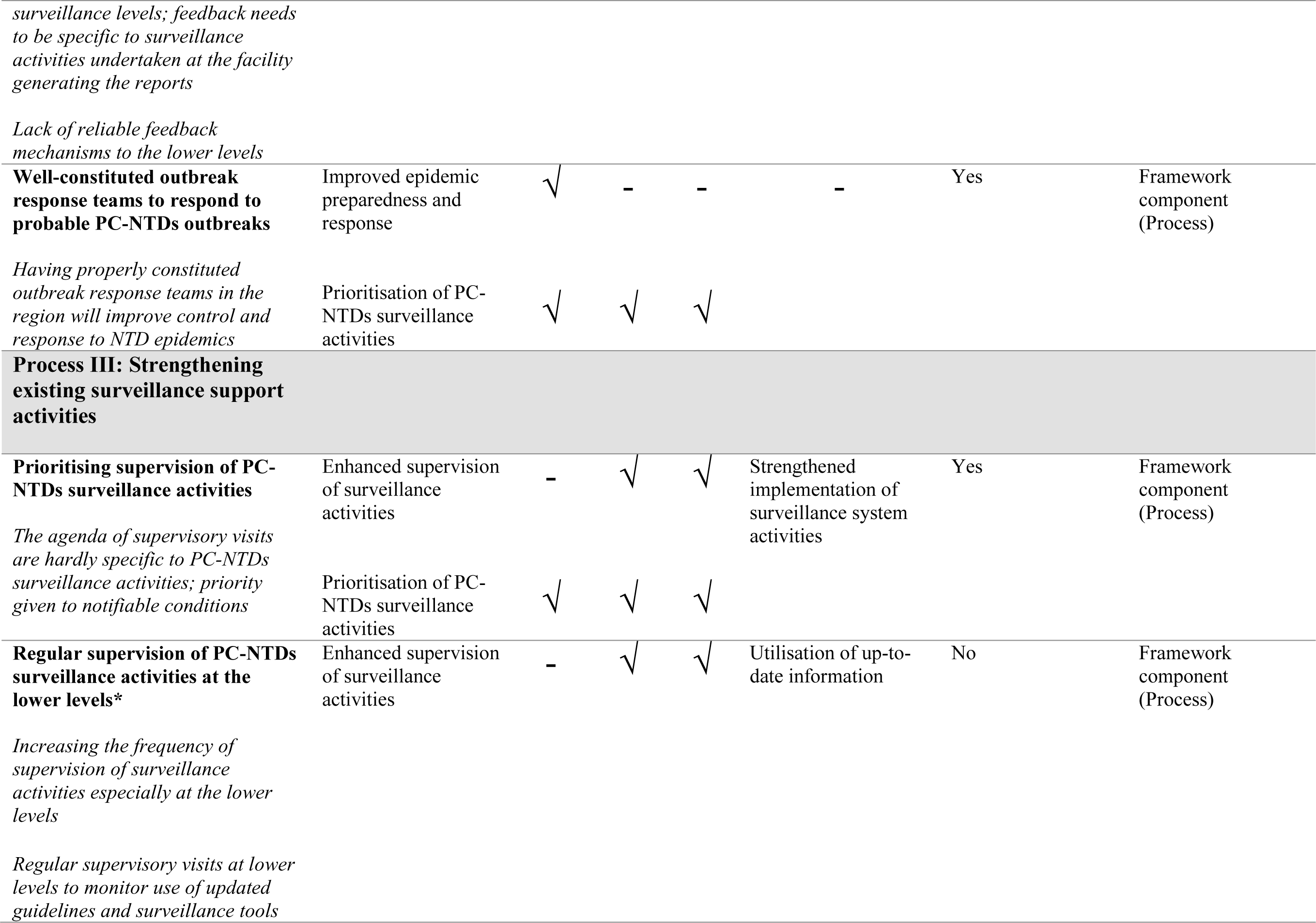
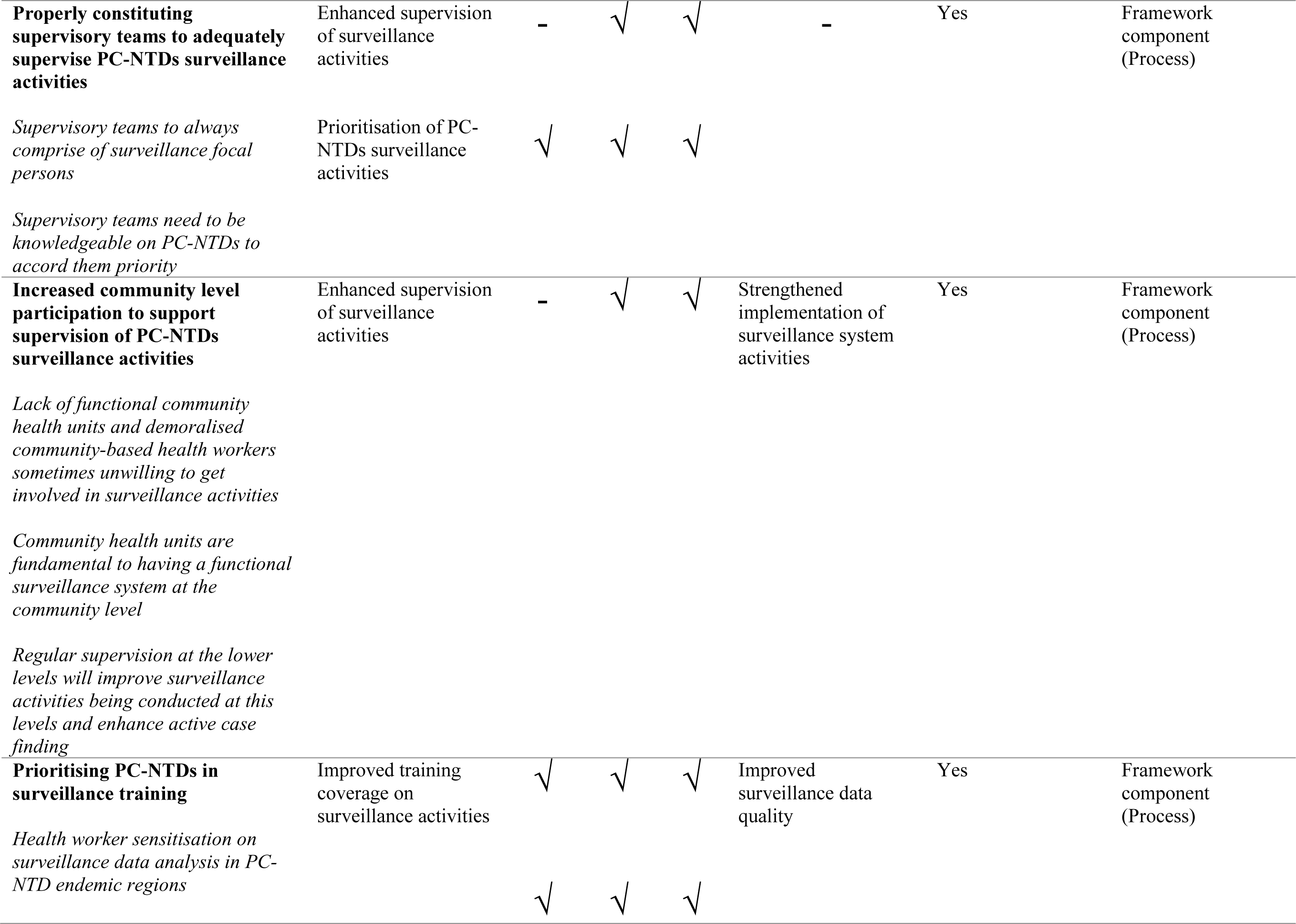
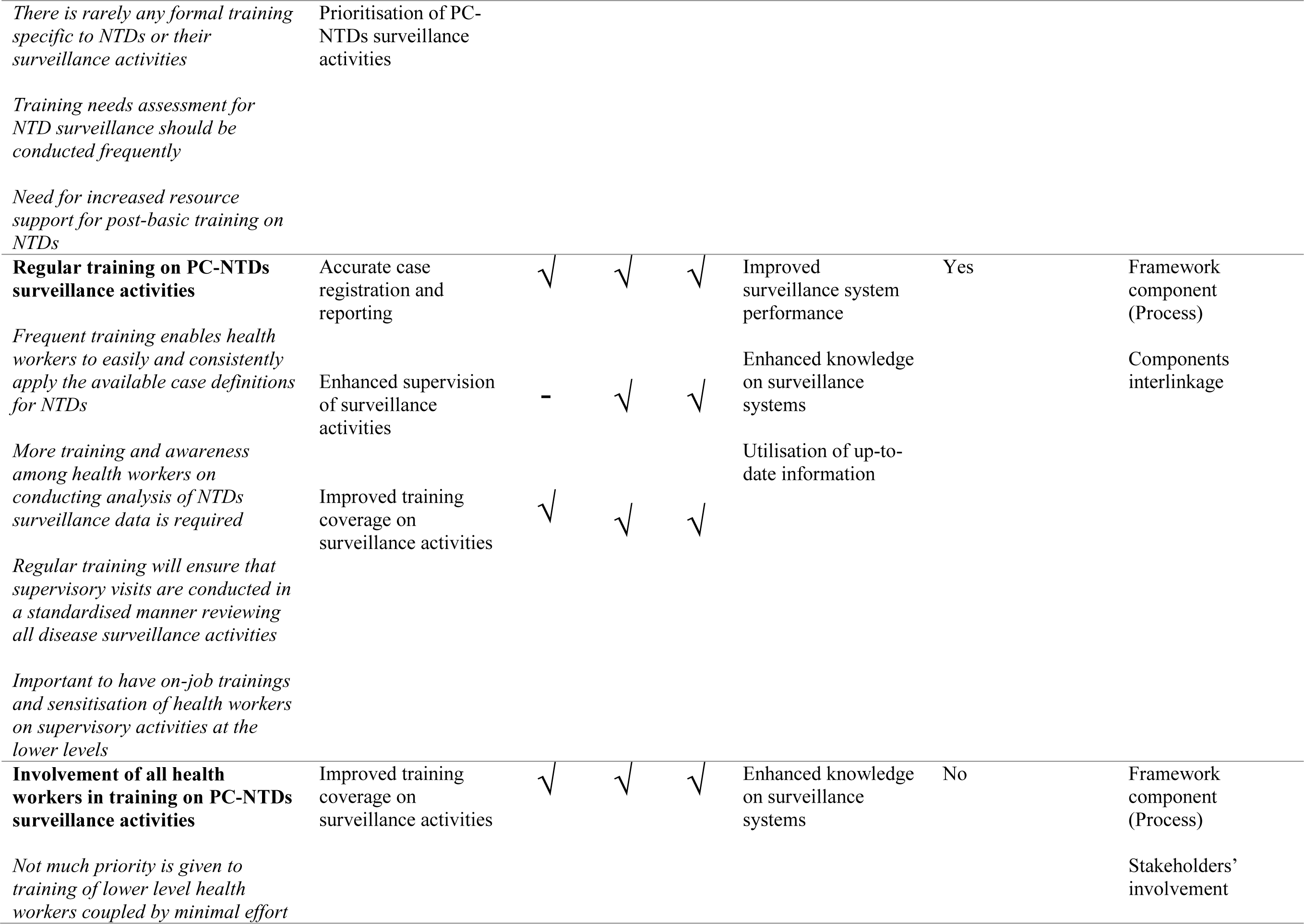
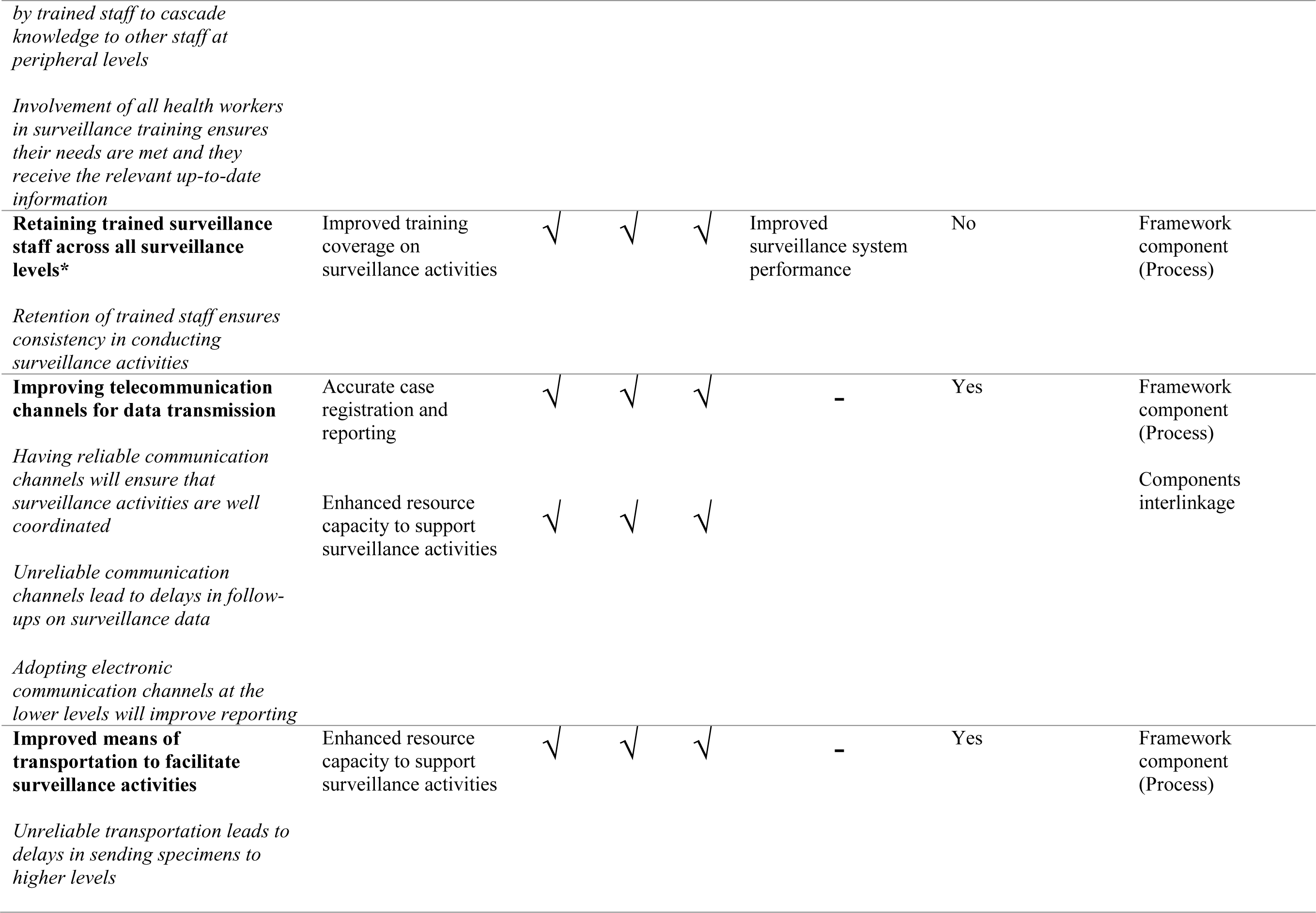
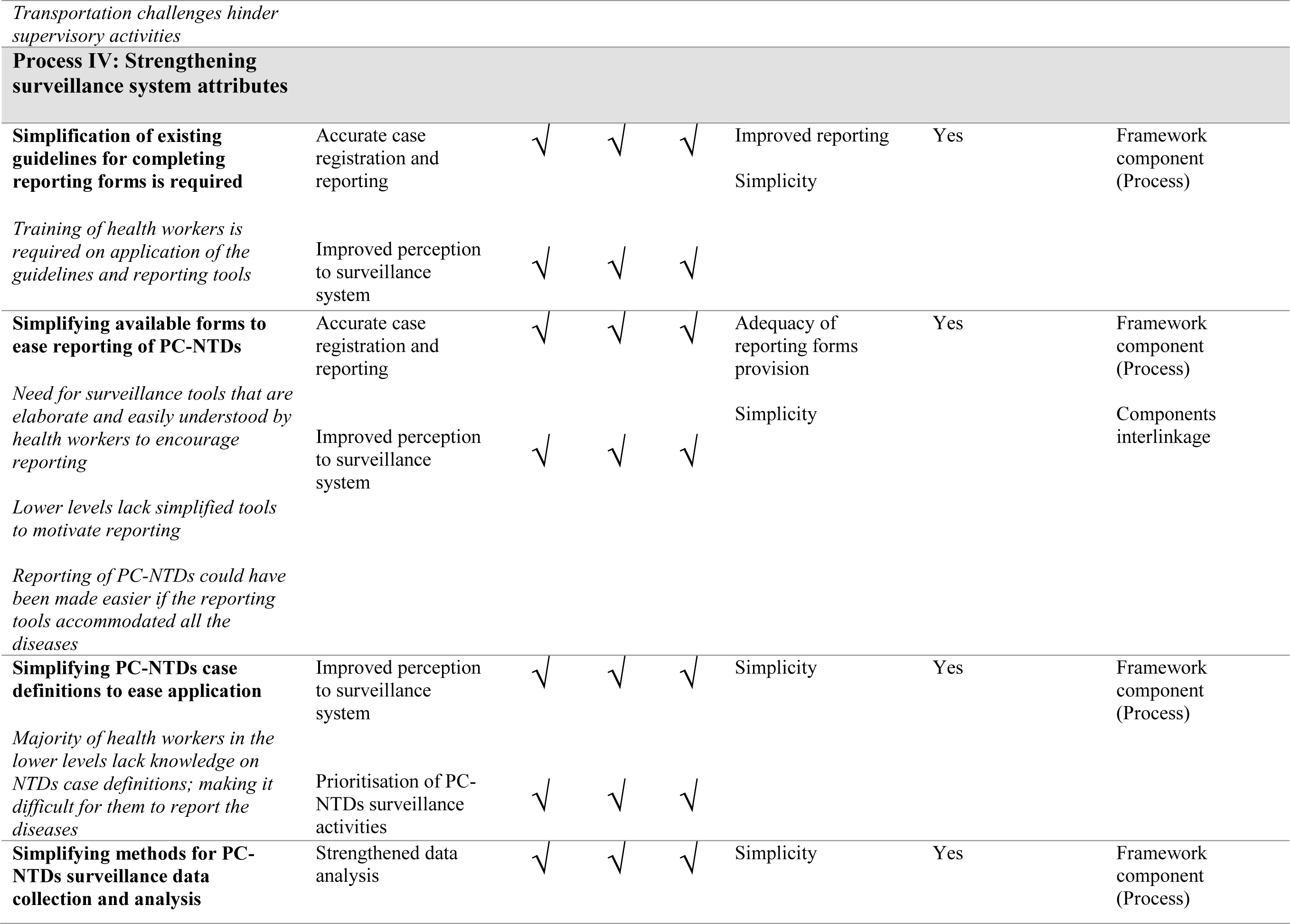
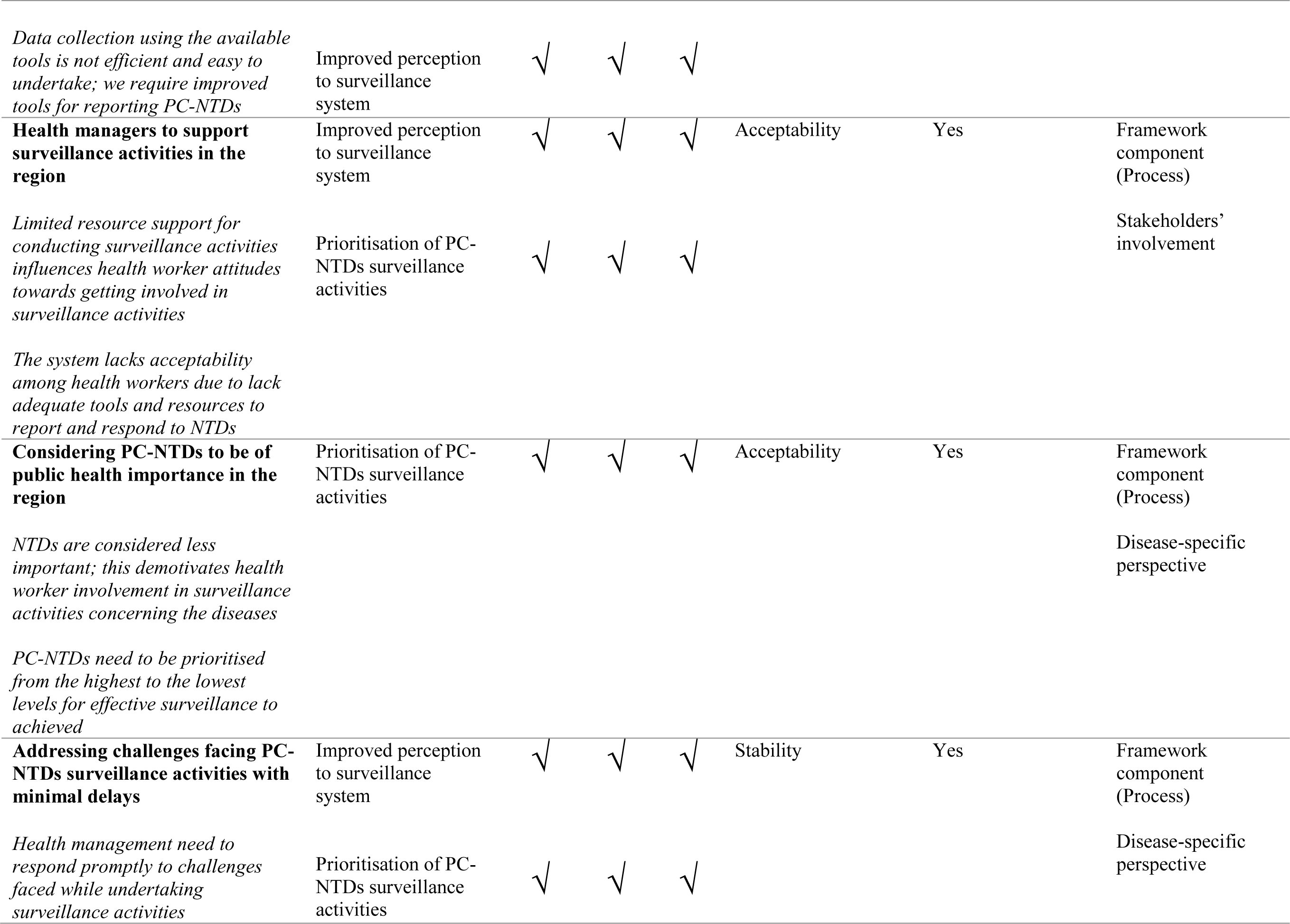
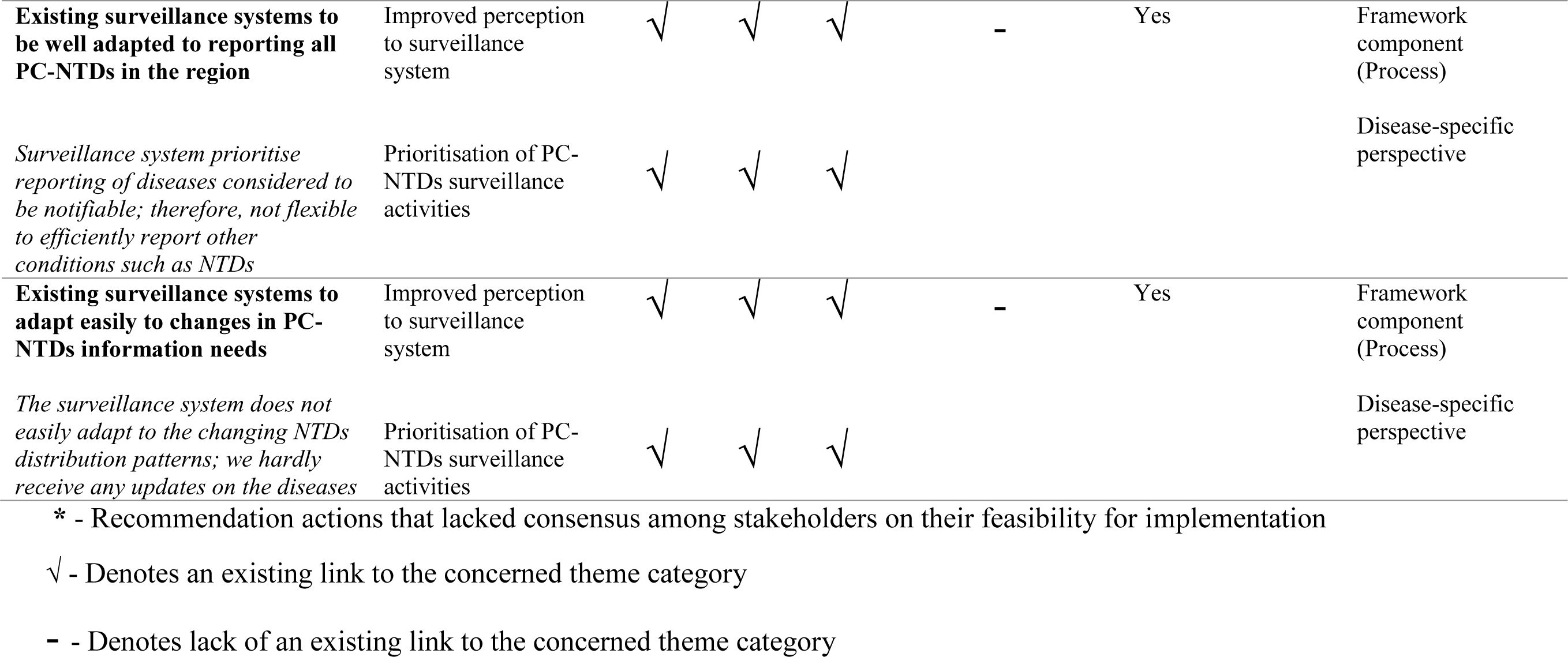
Logical framework component matrix

The first step to developing the proposed framework involved analysing the required inputs. The existing surveillance and response system requires resource mobilisation through injection of inputs to facilitate the planned work in view of NTDs amenable to chemoprophylaxis at the sub- national levels. Therefore, stakeholders’ consensus on the feasible inputs were categorised into two sub-domains with the first being a combination of surveillance tools, equipment and infrastructure including; data analysis tools and equipment such as computers with pre-loaded analysis software, clearly formulated action thresholds, outbreak preparedness and response protocols and guidelines, surveillance activities supervisory schedules, training materials and electronic communication devices. The second sub-domain combined financial, technical and logistical support including; increased funding allocation to support surveillance activities, enhanced training on data analysis, provision of sufficient emergency supplies for outbreak response, reliable transportation and other logistical support.

The second step involved identification of practically feasible processes. The activities were categorised into four sub-domains; (i) strengthening existing surveillance tools, (ii) surveillance core activities, (iii) surveillance support activities, and (iv) surveillance system attributes concerning PC-NTDs. Actionable processes for strengthening the existing surveillance tools included; updating surveillance guidelines and the available standard case definitions currently in use and inclusion of all PC-NTDs in the reporting forms. Processes to strengthen the core activities in view of PC-NTDs included; fully equipping health facility level laboratories, availing updated reporting guidelines, ample time allocation for surveillance reports preparation and submission, enhanced PC-NTDs data analysis, provision of regular and timely feedback, adoption of electronic feedback mechanisms, increased feedback to lower levels and properly constituting outbreak response teams. Additionally, activities to strengthen support activities included; prioritising supervision of PC-NTDs surveillance activities, adequately constituting the supervisory teams, increasing community level involvement in supervisory activities, prioritising and conducting regular PC-NTDs surveillance training, improving telecommunication channels for data transmission and improving access to reliable transportation when undertaking surveillance activities. Lastly, processes to strengthen surveillance attributes regarding PC-NTDs included; simplification of existing reporting guidelines, reporting forms, standard case definitions, and data collection and analysis procedures, increased support for surveillance activities by health managers, consideration of PC-NTDs as conditions of public health importance, addressing surveillance activity challenges with minimum delays, ensuring surveillance systems are well-adapted to reporting all PC-NTDs and easily adapt to changes in information needs.

Consequently, inputs and processes in the planning phase facilitate achieving the intended results with anticipated outputs comprising; accurate case registration and reporting of PC-NTDs, improved case confirmation capacity, strengthened data analysis, improved feedback on surveillance data, improved epidemic preparedness and response, enhanced supervision of surveillance activities, improved training coverage on surveillance activities, enhanced resource capacity to support surveillance activities, improved health worker perceptions towards the surveillance system and prioritisation of PC-NTDs surveillance activities. Resultantly, the outcomes were thematically categorised into three sub-domains, which were linked to the likelihood of achieving either short, medium or long-term outcomes. The short-term outcome alluded to efficient transmission of surveillance data within the existing surveillance system while the medium-term outcome would result to improved surveillance data quality and lastly, the long-term outcome would be early detection and effective response action towards PC-NTDs. The overall impact of the resulting outcomes would be accurate estimation of disease burden, effective identification of disease transmission hotspots and implementation of targeted and cost-effective interventions.

The framework identified the intended results as a by-product of the input and process components making up the initial planned work phase (Figure 2). Consequently, the outputs were linked to one or more anticipated outcomes for strengthening PC-NTDs surveillance and response (Figure 3). First, to achieve the short-term outcome regarding efficient surveillance data transmission and improved data accuracy required enhanced supervision of surveillance activities. Moreover, achieving the medium-term outcome relating to improved surveillance data accuracy and quality relied on strengthened data analysis, improved feedback on surveillance data and enhanced support supervision. Lastly, attaining the long-term outcome of early disease detection and effective response action relied on improved case confirmation capacity, reinforced data analysis and improved epidemic preparedness and response action. Overall, all the three anticipated outcomes – short-term, medium-term and long-term – relied on accurate case registration and reporting, improved training coverage on surveillance, enhanced resource provision, improved health worker perceptions towards the surveillance system and prioritising PC-NTDs surveillance activities. The proposed framework further linked the combination of efficient surveillance data transmission and improved data accuracy and quality to the impact component on accurate estimation of reduced disease burden. Additionally, improved surveillance data accuracy and quality, early disease detection and effective response action were linked to effective identification of disease transmission hotspots. Lastly, the framework identified a link between a combination of short and medium term outcomes to implementation of targeted and cost-effective interventions as an impact component.

**Figure 2.**
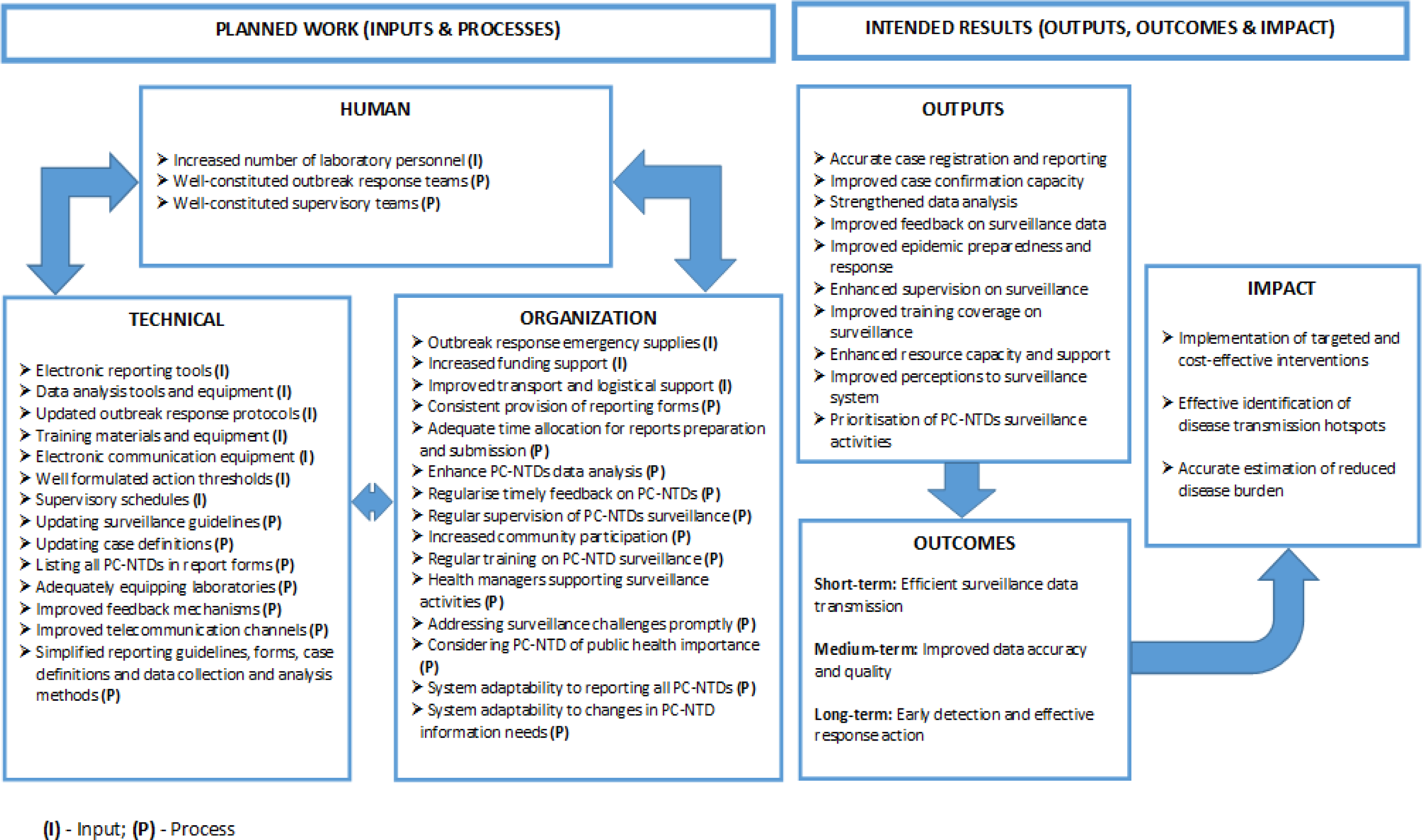
Draft framework for improving PC-NTDs surveillance and response.

**Figure 3.**
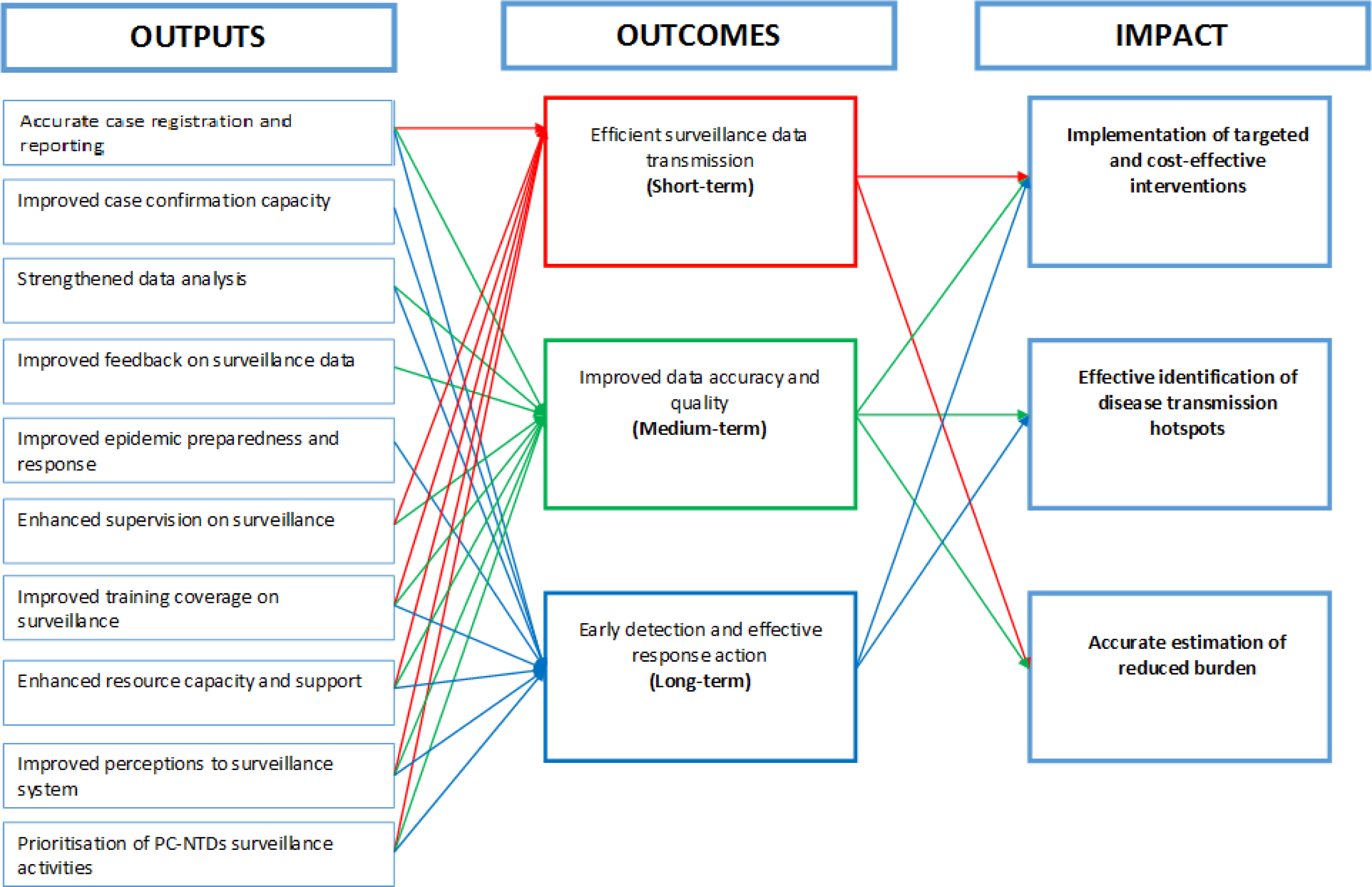
Draft schematic illustration of component interlinkages in the intended results phase.

### Framework validation

The purpose of framework development was to propose a logical approach for strengthening specific surveillance functions at the sub-national level to improve PC-NTDs surveillance and response. Therefore, a multi-phased approach was utilised to validate the proposed framework using: (i) consultative meetings with stakeholders at the sub-national and national levels and (ii) presentation of the draft framework in a conference meeting constituted of NTD researchers and policy experts. The aim of the consultative discussions was to review the draft framework components based on the expertise and experiences of healthcare stakeholders and decision makers. In addition, presentation of the draft framework in a scientific forum was intended to obtain further inputs to improve the framework components and assess scalability and adoptability of the framework especially at the sub-national level based on NTD research experts’ opinions. The validation process presented an opportunity to ascertain the accuracy of information underpinning the draft framework and identified key points for framework refinement (Figure 4). The process allowed participants to comment freely on the framework components in an open forum, which allowed for extensive discussions and drew out diverse stakeholder and expert opinions.

**Figure 4.**
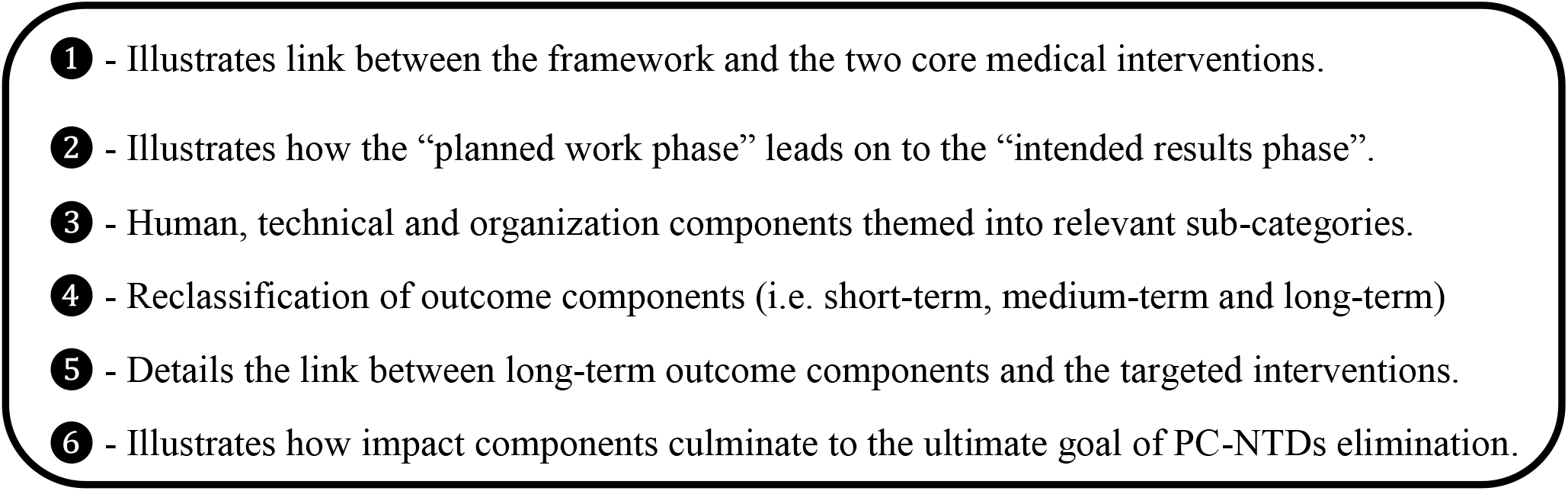
Key points of framework improvement.

### Validation process for the proposed framework

#### Phase I

The stakeholders’ consultative meetings at the sub-national level were conducted in two NTD endemic counties in Kenya. These counties included Baringo and West Pokot, which were purposely selected based on high prevalence of at least three co-endemic PC-NTDs in the regions. Fifteen participants from Baringo (9) and West Pokot (6) counties were enrolled in the validation exercise with representation from both county and sub-county levels. The participants comprised of directors of health, disease surveillance coordinators, epidemiologists, health information and records officers, NTD coordinators, public health officers and other key sub-national health stakeholders. In addition, stakeholder consultations ensued with relevant stakeholders drawn from the national disease surveillance unit (4) and the national NTD programme (3) in Kenya.

The consultative sessions first involved a larger group of participants, whereby the researcher provided a brief background to the framework and described the framework development process. In addition, participants were briefed on the core aim of developing the framework, framework components and their interlinkage. Participants were provided with copies of the draft framework, framework development process including the framework component matrix (Table 1) and a detailed illustration of framework components interlinkage. Subsequently, small group discussions of 3 – 5 participants were formed to further discuss and determine: (1) if the components of the framework required improvements or changes (2) whether the framework presented a logical flow of ideas and (3) the practicability of adopting the framework considering surveillance system capacity and accessible resources at the sub-national level. Broader forums were later reconstituted, involving all participants to further deliberate and reach consensus on resulting outcomes that emanated from the smaller groups.

Resolutions from the consultative sessions were used to improve the draft framework to its final status. The concerned stakeholders reached a number of resolutions on specific framework components (Table 2). First, stakeholders identified the need to establish thematic sub-categories in the planned work phase of the framework. These sub-categories were in line with the broader themes of human, technical and organisational components. Specific sub-themes making up the inputs and processes included human resource management, data management, standards and guidelines, tools and equipment, communication, resource support, surveillance activities management and surveillance system attributes. Secondly, participants identified the need to reclassify the outcome components to derive feasible short-term, medium-term and long-term outcomes. The short-term outcomes were revised to include efficient transmission of surveillance data and improved data accuracy. Medium-term outcomes involved early disease detection and response action and improved surveillance data quality. Lastly, long-term outcomes included; improved implementation of targeted and cost-effective interventions, enhanced identification of disease transmission hotspots, and improved estimation of overall disease burden. Furthermore, participants suggested the need for long-term outcome components to be linked to target interventions. For instance, enhanced identification of disease transmission hotspots was a desired long-term outcome, which was linked to intensified case finding (CF). Thirdly, participants suggested the need to revise impact components to signify sustainable efforts resulting from the long-term outcomes towards achieving the ultimate goal of PC-NTDs elimination. Therefore, the overall impact components were revised to include reduced costs for implementing PC interventions, halted disease transmission and reduced disease burden relating to PC-NTDs.

**Table 2.**
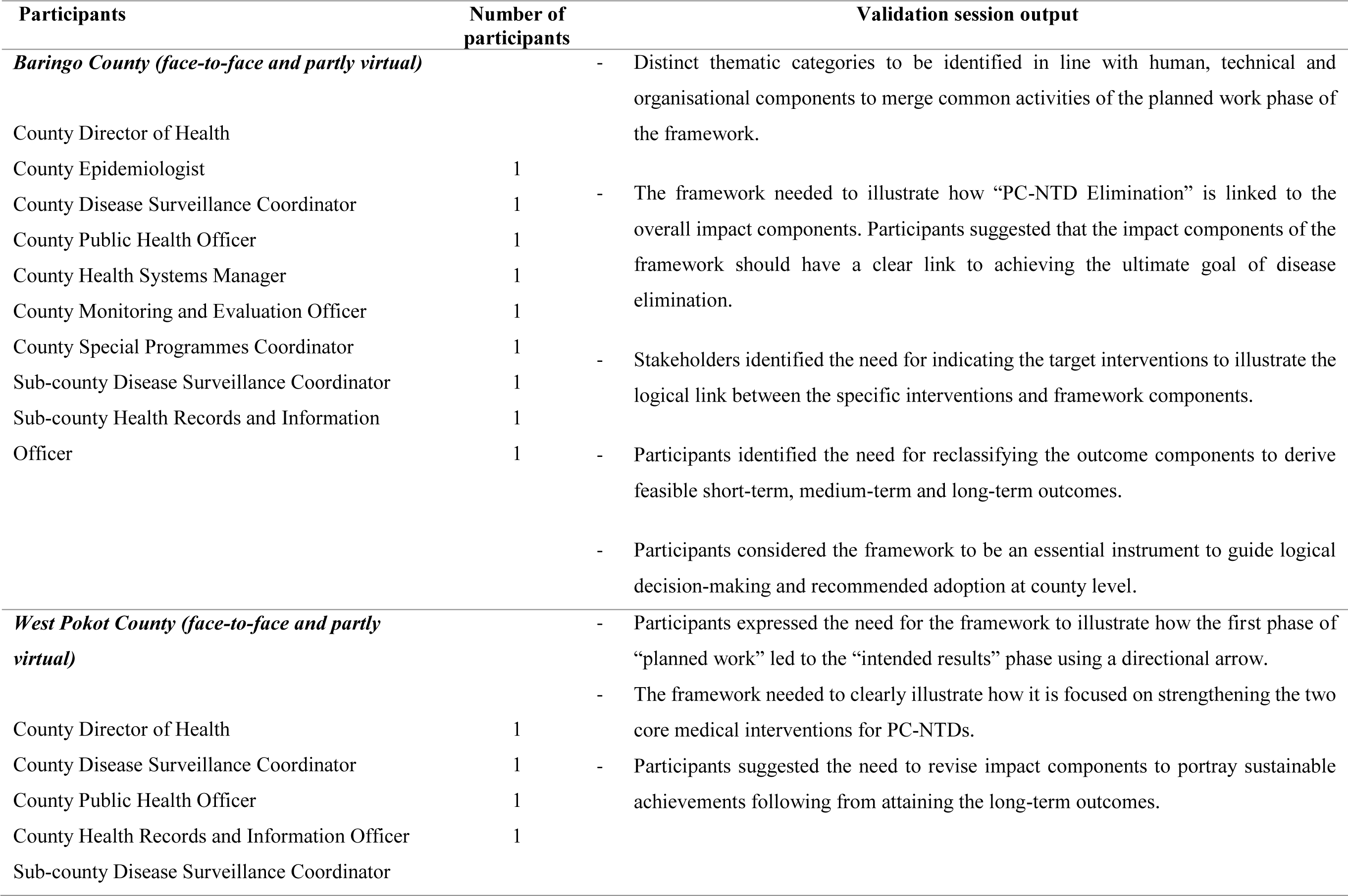
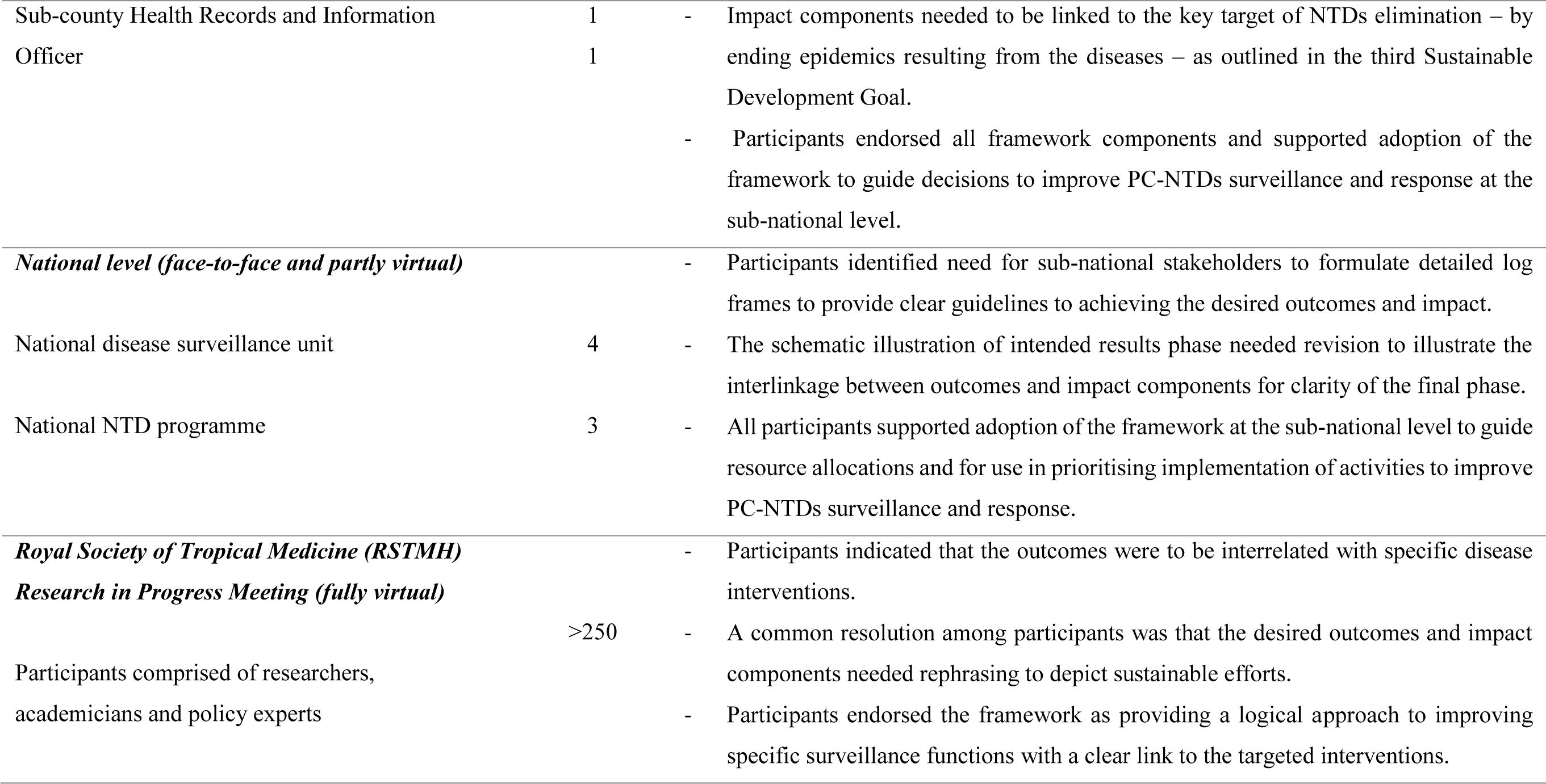
Framework validation resolutions

Fourthly, stakeholders indicated that the framework needed to illustrate the link between specific medical interventions associated with framework components. Therefore, the framework was shown to target improvement of PC and intensified disease management (IDM) interventions. Moreover, participants suggested the need for impact components of the framework to be linked to the ultimate goal of disease elimination as outlined in the NTD roadmap and in achieving the third SDG [68, 69]. Further resolutions alluded to illustrating how the “planned work” phase led on to the “intended results” phase using a directional arrow. Lastly, participants at the sub-national level endorsed the framework as identifying feasible actions for strengthening specific surveillance functions in relation to PC-NTDs.

On the other hand, participants involved in the framework validation process at national level recommended that the schematic illustration of the intended results phase focus on illustrating the interlinkage between desired outcomes and impact components. Moreover, participants recognised need for supplementing the logical framework with detailed log frames for the desired impacts to be realised. The log frames for achieving the three distinct impact components constituted objectively verifiable indicators, sources of information, inputs (resources) and relevant assumptions (Table 3-5). Log frames formulated through consultations with concerned stakeholders depicted that the overall impact of disease elimination was achievable through an integrated approach. This meant consolidating various inputs and processes at the planned work phase to achieve the intended results and outcomes. Essentially, stakeholders agreed that the framework offered a logical approach to improve PC-NTD surveillance and response from the initial planned work phase to achieving the intended results and the ultimate goal of disease elimination.

**Table 3.** Log frame 1

| Objectives |  | Indicators of achievement | Means of verification | Important assumptions |
| --- | --- | --- | --- | --- |
| Goal (Impact) | Reduced costs for treatment interventions | Decreased number of regions and population requiring treatment | -Survey reports<br>-County health reports<br>-Community Drug Distributors (CDDs) reports | -Political goodwill<br>-Donor and partner support<br>-Community perceptions and participation |
| Purpose (Outcome) | Implementation of targeted and cost effective interventions<br>**(Target intervention: PC)** | -Percentage of HFs with improved reporting rates<br>-Percentage of HFs registering and reporting PC-NTDs cases<br>-Percentage of Sub-counties implementing PC-interventions informed by surveillance data | -Survey reports<br>-County health reports<br>-Record reviews<br>-DHIS2 | -Availability and adequacy of surveillance tools<br>-Submission of complete and timely reports |
| Outputs | -Accurate case registration and reporting<br>-Improved case confirmation capacity<br>-Strengthened data analysis<br>-Improved feedback on surveillance data<br>-Improved epidemic preparedness and response<br>-Enhance supervision on surveillance<br>-Improved training coverage on surveillance<br>-Enhanced resource capacity and support<br>-Improved perceptions to surveillance system<br>-Prioritisation of PC-NTDs surveillance activities | -Proportion of SUs registering and reporting PC-NTDs data<br>-Percentage of HFs with functional laboratories<br>-Proportion of HFs analysing PC-NTDs data<br>-Proportion of SUs providing feedback on PC-NTDs<br>-Proportion of SUs with adequate outbreak response supplies<br>-Proportion of SUs supervised on PC-NTDs surveillance activities<br>-Proportion of SUs with health workers trained on PC-NTDs surveillance<br>-Proportion of SU with adequate surveillance resources<br>-Proportion of SU with health personnel willing to be involved in PC-NTDs surveillance activities<br>-Proportion of SU with PC-NTDs yearly surveillance plans | -Survey reports<br>-County health reports<br>-Record reviews<br>-DHIS2 | -Infrastructural and technological improvements<br>-Availability and adequacy of surveillance tools and guidelines<br>-Continuous training of healthcare workers and provision of adequate training resources |
| Activities (inputs & processes) | Technical and Organisational | -Data management (i.e. electronic reporting and data analysis tools)<br>-Standards and guidelines (i.e. simplified reporting guidelines, report forms, standard case definitions and data collection and analysis)<br>-Communication (i.e. electronic equipment and improved communication channels)<br>-Resource support (i.e. increased funding and provision of reporting forms) | -Survey reports<br>-County health reports<br>-County budgetary allocation reports | -Infrastructural and technological improvements<br>-Availability and adequacy of funding support<br>-Prioritising funding for PC-NTDs surveillance activities |

**DHIS2:** District Health Information System, **HFs:** Health Facilities; **SUs:** Surveillance Units (surveillance levels within sub-national structures); **PC:** Preventive Chemoprophylaxis; **PC-NTDs:** Preventive Chemotherapy-targeted Neglected Tropical Diseases

**Table 4.** Log frame 2

| **Objectives** |  | **Indicators of achievement** | **Means of verification** | **Important assumptions** |
| --- | --- | --- | --- | --- |
| Goal (Impact) | Halted disease transmission | Reduced PC-NTDs transmission hotspots | -Survey reports (i.e. TAS)<br>-County health reports | -Political goodwill<br>-County government support<br>-Donor and partner support<br>-Complete disease mapping<br>-Community perceptions and participation |
| Purpose (Outcome) | Enhanced identification of disease transmission hotspots<br>**(Target Interventions: CF)** | -Percentage of SUs registering and reporting PC-NTDs data<br>-Percentage of SUs notifying on PC-NTDs cases | -DHIS2<br>-Record reviews<br>-Feedback bulletins | -Availability and adequacy of surveillance tools and guidelines |
| Outputs | -Accurate case registration and reporting<br>-Improved case confirmation capacity<br>-Strengthened data analysis<br>-Improved feedback on surveillance data<br>-Improved epidemic preparedness and response<br>-Enhance supervision on surveillance<br>-Improved training coverage on surveillance<br>-Enhanced resource capacity and support<br>-Improved perceptions to surveillance system<br>-Prioritisation of PC-NTDs surveillance activities | -Proportion of SUs registering and reporting PC-NTDs data<br>-Percentage of HFs with functional laboratories<br>-Proportion of HFs analysing PC-NTDs data<br>-Proportion of SUs providing feedback on PC-NTDs<br>-Proportion of SUs with adequate outbreak response supplies<br>-Proportion of SUs supervised on PC-NTDs surveillance activities<br>-Proportion of SUs with health workers trained on PC-NTDs surveillance<br>-Proportion of SUs with adequate surveillance resources<br>-Proportion of SUs with health personnel willing to be involved in PC-NTDs surveillance activities<br>-Proportion of SUs with PC-NTDs yearly surveillance plans | -Survey reports<br>-County health reports<br>-DHIS2<br>-Record reviews | -Infrastructural and technological improvements<br>-Continuous training of healthcare workers and provision of adequate training resources |
| Activities (inputs and processes) | Human, Technical and Organisational | -Human resource management (i.e. outbreak response teams and laboratory personnel)<br>-Data management (i.e. provision of electronic reporting tools and listing all PC-NTDs in reporting forms)<br>-Standards and guidelines (i.e. updated case definitions and surveillance guidelines)<br>-Tools and equipment (i.e. adequately equipped laboratories)<br>-Resource support (i.e. increased funding support)<br>-Surveillance activities management (i.e. enhanced data analysis and increased community participation)<br>-Surveillance system attributes (i.e. considering PC-NTDs of public health importance) | -Survey reports<br>-County Health reports<br>-County Health budgetary allocation reports | -Availability and adequacy of funding support<br>-Prioritising funding for PC-NTDs surveillance activities |

**CF:** Case Finding; **DHIS2:** District Health Information System, **HFs:** Health Facility; **SUs:** Surveillance Units (surveillance levels within sub-national structures); **PC-NTDs:** Preventive Chemotherapy-targeted Neglected Tropical Diseases; **TAS:** Transmission Assessment Surveys

**Table 5.** Log frame 3

| Objectives |  | Indicators of achievement | Means of verification | Important assumptions |
| --- | --- | --- | --- | --- |
| Goal (Impact) | Reduced disease burden related to PC-NTDs | Decreased number of cases requiring IDM | -Survey reports<br>-County health reports | -Political goodwill<br>-County government support<br>-Donor and partner support<br>-Community perceptions and participation |
| Purpose (Outcome) | Improved estimation of overall disease burden<br>**(Target intervention: IDM)** | -Percentage of SUs submitting complete reports on PC-NTDs data<br>-Percentage of SUs with improved reporting rates | -Record reviews<br>-DHIS2 | -Availability and adequacy of surveillance tools and guidelines |
| Outputs | -Accurate case registration and reporting<br>-Strengthened data analysis<br>-Improved feedback on surveillance data<br>-Enhance supervision on surveillance<br>-Improved training coverage on surveillance<br>-Enhanced resource capacity and support<br>-Improved perceptions to surveillance system<br>-Prioritisation of PC-NTDs surveillance activities | -Proportion of HFs registering and reporting PC-NTDs data<br>-Proportion of HFs analysing data<br>-Proportion of SUs providing feedback<br>-Proportion of HFs supervised on NTDs<br>-Proportion of SUs with trained HCWs<br>-Proportion of SUs with adequate resources<br>-Proportion of SUs with health personnel willing to be involved in PC-NTD surveillance activities<br>-Percentage of sub-counties with PC-NTD surveillance plans | -Record reviews<br>-DHIS2<br>-Survey reports<br>-County health reports<br>-Feedback bulletins | -Complete and timely reporting<br>-Regular feedback and use of appropriate feedback mechanisms<br>-Continuous training of HCWs |
| Activities (inputs and processes) | Human, Technical and Organisational | -Human resource management (i.e. well constituted supervisory teams)<br>-Data management (i.e. electronic reporting and data analysis tools)<br>-Standards and guidelines (i.e. updated surveillance guidelines)<br>-Tools and equipment (i.e. training materials and equipment and supervisory schedules)<br>-Communication (i.e. electronic communication channels and improved feedback mechanisms)<br>-Resource support (i.e. increased funding support and consistent provision of report forms)<br>-Strengthened management of surveillance activities (i.e. enhanced data analysis, regular feedback and supervision and increased community participation)<br>-Surveillance system attributes (i.e. consideration of PC-NTDs on public health importance, health management support and system adaptability) | -Survey reports<br>-County health reports<br>-County Health budgetary allocation reports | -Availability and adequacy of funding support<br>-Prioritising funding for PC-NTDs surveillance activities |

**DHIS2:** District Health Information System, **HCWs:** Healthcare Workers, **HFs:** Health Facilities, **IDM:** Intensified Disease Management; **SUs:** Surveillance Units (surveillance levels within sub-national structures); **PC:** Preventive Chemoprophylaxis; **PC-NTDs:** Preventive Chemotherapy-targeted Neglected Tropical Diseases

#### Phase II

The framework development process was further presented in a virtual conference meeting in the form of a research poster. The forum was partly composed of breakout sessions that enabled extensive discussions of the framework components with various NTD researchers and policy experts. Participants identified the need for interrelating the desired outcomes with the concerned disease interventions, similar to stakeholder resolutions in the first phase. They envisaged that such an interrelation would provide a clear link between implementing specific control interventions and achieving the overall goals. Additionally, participants recommended that the impact components be rephrased to portray “sustained” control efforts even post-elimination. Lastly, participants reached a common consensus on the relevance and feasibility of adopting the proposed framework at sub-national levels. They further agreed that the framework provided a logical approach to guide stakeholders’ actions to achieve improved PC-NTDs surveillance and response in endemic settings (Figure 5). Stakeholder consultations in both phases aided in identifying critical interlinkages between the long-term outcomes and impact components in various combinations (Figure 6). First, improved implementation of targeted and cost-effective PC interventions, in addition to enhancing identification of disease transmission hotspots through active CF would have an overall impact of reducing costs for implementing treatment interventions. Secondly, identification of disease transmission hotspots and improved estimation of overall disease burden would inform effective implementation of PC interventions and intensify disease management, which would contribute to halted disease transmission. Finally, a combination of all long-term outcomes on implementation of targeted PC interventions, identification of disease transmission hotspots through active CF and improved estimation of overall disease burden to inform IDM would achieve reduced disease burden relating PC-NTDs.

**Figure 5.**
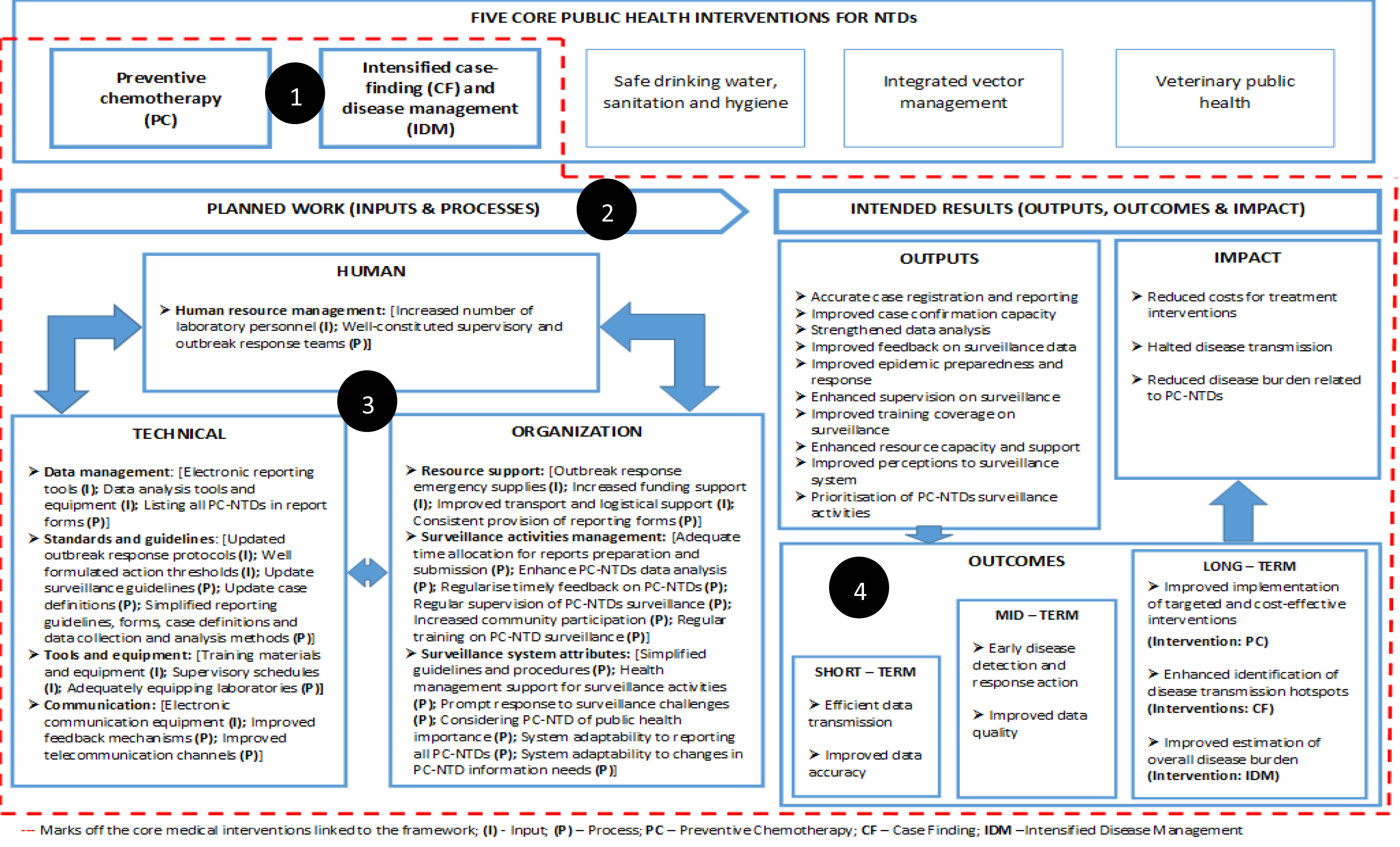
Final validated framework for improving PC-NTDs surveillance and response.

**Figure 6.**
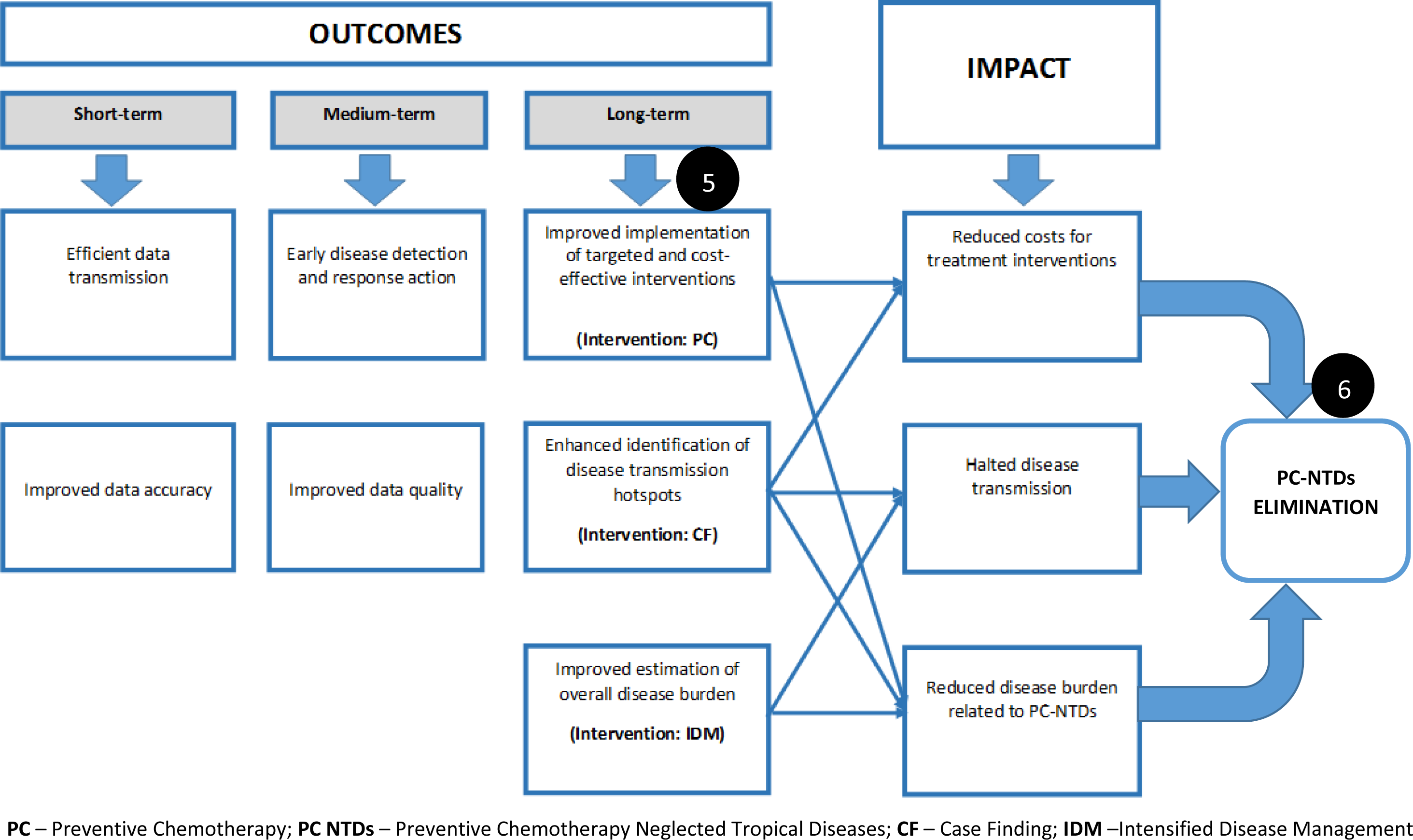
Final schematic illustration of component interlinkages in the intended results phase.

The overall impact of reduced costs for treatment interventions, halted disease transmission and reduced disease burden contribute towards achieving sustainable PC-NTDs elimination.

## Discussion

We developed a logical framework for guiding implementation of recommendations to improve surveillance and response to NTDs amenable to chemoprophylaxis. The framework was grounded on existing conceptual frameworks and adopted a stakeholder-oriented approach to determine the framework components and their interlinkage [19–21, 65]. The feasible actions identified in the third phase of framework development were assessed and categorised as either constituting input or process components. The outputs, outcomes and impact components were thematically derived from reviewed literature in the first phase and analysis of participants’ open-responses in the second and third phases. Recommendations drawn from Phases I, II and III were incorporated into a logical component matrix to assess interdependence between the inputs, processes, outputs, outcomes and overall impact. Furthermore, we utilised the W. K. Kellogg Basic Logic Model to derive key constructs and themes as described by concerned health stakeholders [64]. The logic model provides a systematic and visual course of action to recognise the link between resources or inputs, planned activities and the intended results [64].

Initial emphasis was on input and process indicators that form the core of the planned surveillance work following which priority shifts methodically to the outcome and impact indicators. Surveillance and response systems, although fundamental, they perform sub-optimally in the case of PC-NTDs. Collation of previous and present information on NTDs through reviewing both published and grey literature, analysis of relevant health data and seeking local stakeholders views form the basis for achieving integrated NTDs control [70]. Hence, the validated framework considered these key features to contribute towards efforts to achieve an integrated approach for PC-NTDs control through strengthened disease surveillance systems. Implementation of interventions involving multiple diseases is imperative and cost-effective especially in NTD endemic regions as opposed to parallel disease-specific strategies [71]. The WHO categories twenty diseases and conditions to constitute NTDs with two distinct control measures; those diseases that are amenable to preventive chemoprophylaxis and those tackled through case management (CM) [7, 8]. The focus of the current framework was on NTDs mainly controlled through PC interventions including lymphatic filariasis (LF), schistosomiasis, trachoma and soil transmitted helminth (STH) infections. Nonetheless, effective control of some of the PC-NTDs including LF and trachoma requires IDM [8, 25]. Furthermore, WHO recommends five key strategies for prevention, control, elimination and eradication of NTDs [71, 72]. These strategies include PC and IDM, which are the core medical interventions for curbing NTDs transmission. The other cross-cutting strategies include vector and intermediate host control, veterinary public health and provision of safe water, sanitation and hygiene [72]. Therefore, the present framework intended to link strengthened surveillance and response systems to the two core medical intervention strategies to achieve effective PC-NTDs control. Noteworthy, disease surveillance systems provide essential tools for verification and improvement of interventions during implementation [72].

The first phase of framework development involved conducting a systematic literature review of previous surveillance assessment studies in the African region to assess performance of surveillance functions based on health workers’ perspectives. The review revealed a dearth in surveillance assessment studies conducted in African countries with a focus on NTDs, which informed the second phase involving primary data surveys undertaken in an NTD endemic setting. An assessment of surveillance core, support and attribute functions regarding PC-NTDs targeted for control and elimination identified critical recommendations to improve their surveillance and response in Kenya. The third phase was aimed at determining the importance and feasibility of implementing the actions to improve PC-NTDs surveillance and response as perceived by healthcare workers in the second phase and generalised recommendations retrieved from the systematic literature review in the first phase. Consequently, the proposed framework components were informed by findings emerging from the three phases and a fourth phase involving a review of relevant conceptual frameworks formed the base for developing the current framework. Furthermore, existing strategic plans focused on NTDs control and elimination formed the blueprint for framework development. The National Breaking Transmission Strategy (BTS) for NTDs in Kenya, which is founded on the WHO roadmap for NTDs identifies pertinent strategies to achieve effective NTDs control [31, 72]. A key strategic objective of the BTS is focused on strengthening surveillance systems, which is also in line with the objective on strengthening information systems for evidence-based action as stipulated by the Expanded Special Project for Elimination of NTDs (ESPEN) [31, 73]. Additionally, the forth-strategic objective of the Second Kenya National Strategic Plan for Control of NTDs 2016-2020 aimed to enhance NTDs monitoring and surveillance activities [30]. Therefore, the current framework is geared towards meeting these key strategic objectives to improve the overall surveillance and response system with regard to NTDs. The framework further looks to contribute towards efforts aimed at accelerating programmatic actions to control and eliminate NTDs as outlined in the recently launched Roadmap for NTDs 2021-2030 [68]. Essentially, the framework is inclined to improving the efficiency of medical interventions relating to implementation of PC, which is considered the safest and most effective intervention for controlling PC-NTDs. In addition, achieving NTDs- related morbidity reduction requires IDM, which involves early disease detection through active CF, treatment and clinical management critical to NTDs control and elimination [8].

The framework provided a logical approach constituting of interlinked inputs, processes, outputs, outcomes and impact factors. A technical input incorporated in the framework was the provision of up-to-date surveillance guidelines, which corresponds to meeting NTD elimination targets by way of providing standardised guidelines essential for systematic collection of population-based data [74]. Furthermore, utilisation of WHO-approved guidelines will aid disease mapping, monitoring and post-MDA surveillance approaches [74]. On the other hand, implementation of integrated and targeted PC interventions are largely impeded by the lack of complete data [70]. Hence, improving the existing reporting forms to ensure accurate and complete reporting of PC-NTDs is justified. There are no standardised methods for collecting PC-NTDs morbidity data except through baseline surveys during MDA campaigns [75]. Therefore, morbidity data collected while concurrently conducting MDA usually lack quality and accuracy [76]. This affects effective implementation of NTD-targeted morbidity management strategies that are dependent on rapid generation of information on the disease burden in endemic regions [75]. Furthermore, countries requiring validation and eventual certification as having eliminated specific NTDs need to accurately estimate the number of cases in endemic regions at the implementation unit level and monitor morbidity management and disability prevention (MMDP) [76]. This requirement corresponds to the framework’s long-term outcome on improved estimation of overall PC-NTD burden.

Core to developing the framework was to involve healthcare stakeholders at the sub-national level, responsible for overseeing disease surveillance activities to identify the feasible actions to improve surveillance activities relating to PC-NTDs. Notably, a key challenge in the PC-NTDs control and elimination agenda in Kenya is the minimal national and sub-national ownership of planned actions with most funding support for implementing interventions coming from development partners. In addition, limited involvement of local health stakeholders in NTDs control activities presents a pivotal sustainability challenge [31]. However, the validated framework components were identified based on convergence of opinion among various stakeholders. Stakeholder involvement and consensus is vital in guiding HIS strengthening and informing key healthcare decisions [18, 19]. The framework further identified the need for adequate surveillance technical tools, regular training of health workers, increased community participation in surveillance activities and the need for health management levels within sub-national structures to support surveillance activities by providing sufficient resources. Comparably, a critical outcome from the ‘First Forum on Surveillance-Response System Leading to Tropical Disease Elimination’ held on June 16-18, 2012 in Shanghai, identified key features influencing effective surveillance and elimination of NTDs [77]. These factors alluded to political will and good governance, commitment to utilise available resources to support disease elimination efforts, an established organisational and technical infrastructure with a competent health workforce, reliable healthcare services delivery and community involvement [77, 78]. Therefore, critical elements of the validated framework are in line with meeting key targets to achieve NTDs elimination. Furthermore, increased knowledge on co-endemic NTDs may improve prompt case detection of multiple diseases that would eventually inform integrated management [25]. However, effective NTD training can only be achieved when disease-specific materials are utilised rather than adopting integrated health education approaches using complex guidelines [70]. This is consistent with the current framework process component on instituting regular training specific to PC-NTDs surveillance.

On the other hand, encouraging involvement of the community levels in surveillance activities is deemed to increase ownership and sustainability of the processes. This corresponds to efforts to empower communities living in NTD endemic regions through their participation in control interventions and formalizing the roles of community health workers [8, 67]. Minimal training at the community level on lay reporting and identifying NTDs with visible manifestations through use of the local terms and simple examinations contributes to valid estimation of disease prevalence in endemic regions [79]. Further, active CF at the peripheral level warrants reinforcement of community and health workers knowledge on NTDs through increased sensitisation [25]. Community health structures are an inherent part of health systems in developing countries especially regarding disease surveillance-related activities and efforts to achieve disease control. This is commensurate with the important assumption detailed in the log frames on increased community involvement [75]. Therefore, improved resource allocation at the sub-national level will support capacity building and training for community-based health workers resulting to strengthened community-level structures [31]. Effective surveillance systems may contribute immensely to active detection of symptomatic cases at the lower levels. However, measurement of NTDs burden may be hindered in a majority of endemic settings with substantial proportions of asymptomatic cases similar to challenges facing malaria elimination programmes [80].

The framework’s input and process components were broadly categorised into human, technical and organisational aspects. This was partly informed by the PRISM framework that clearly describes the interconnectedness between technical, behavioral and organisational factors [20]. Furthermore, the proposed framework adopted the HOT-fit concept that considers development of HIS according to an organisational plan of action while concurrently managing information technologies based on organisational needs [21]. The model considers human, organisation and technology as essential components of HIS, which influence information and service quality, system utilisation, user satisfaction, organisational environment and the net benefits [21]. Comparably, the validated framework combined human, technical and organisational input and process components to achieve the intended results of improved data quality, improved system perceptions, enhanced surveillance system use and achieving the overall impact on improved disease control. The framework further depicts a linkage between public health surveillance core, support and attribute functions and public health actions including implementation of targeted and cost-effective interventions, identification of disease transmission hotspots and accurate disease burden estimation. This compares closely to the concept of linking public health surveillance activities to public health actions – acute or planned response – through a series of process-oriented measures to achieve the intended outputs and outcomes [17, 18]. Moreover, in the planned work phase of the current framework, having adequate epidemic preparedness and response emergency supplies and well-constituted outbreak response teams were identified as critical inputs and processes respectively. Achieving improved epidemic detection as a key output through processes involving data collection, analysis and epidemiologic investigations were synonymous with the framework for evaluating public health surveillance systems for early outbreak detection [34]. Likewise, ending NTD epidemics is a crucial target to achieving the third SDG [69].

The current framework identified a critical interrelation between reduced PC-NTDs burden and implementation of targeted and cost-effective treatment interventions coupled with identification of disease transmission hotspots. Comparably, to achieve NTDs elimination goals there has to be an intentional shift towards implementing PC strategies to not only attain conservative morbidity control but towards progressive disease transmission interruption [81]. Furthermore, implementation of cost-effective interventions rely on a shift from MDA to limiting the interventions to specific at-risk population groups [81]. Albeit the substantial progress so far achieved to control NTDs through cost-effective interventions, the remaining challenge of instituting adequate monitoring and surveillance efforts hinder sustained disease control and elimination [82]. In particular, disease surveillance is a dynamic process that involves collection of a critical set of data to inform public health actions based on specific thresholds. This is exemplified at the disease elimination stage, whereby an effective surveillance system would enable prompt detection of persisting or re-emerging transmission foci, detect instances of drug resistance and the identification of low transmission regions [78]. Therefore, surveillance systems aid in the identification of disease transmission foci to inform targeted response actions in preference to mass interventional campaigns, which may otherwise result to misuse of scarce resources. In addition, continued mass treatment interventions may exert pressure on the disease causative agents resulting to drug resistance [83]. Therefore, targeted disease control strategies and implementation of cost-effective interventions in areas recognised as disease transmission hotspots are essential to attaining eventual PC-NTDs elimination. The validated framework further identifies the long-term component on improved estimation of overall disease burden as an enabler for IDM. For instance, effective surveillance systems could pinpoint regions with a high burden of progressed forms of trachoma and LF, which may inform effective implementation of trichiasis surgery to prevent corneal scarring and hydrocele surgery to prevent male genital deformities respectively [84].

Framework validation process identified opportunities for strengthening specific framework components. Stakeholder resolutions provided insights on the scalability, adoptability and feasibility of the framework to guide decisions for improving PC-NTDs surveillance and response at the sub-national level. Additionally, prior consensus statements in the Delphi survey identified actionable recommendations according to concerned health stakeholders, which informed components constituting the current framework. The log frames guided improved planning, implementation and management of activities to achieve desired goals [85]. They enabled structuring of framework elements – inputs, processes, outputs, outcomes and impact – to highlight the logical interlinkages [85]. In addition, log frames provided answers to critical questions at every phase of implementation; (i) if activities are implemented, will outputs be produced?, (ii) if outputs are produced, will specific outcomes result?, (iii) if outcomes result, will the objectives be achieved?, (iv) will achieving the objectives contribute to the desired impact or the larger goal?. Therefore, these questions were answered by way of a four-by-four matrix, on one side providing a hierarchy of objectives including; (i) goal (overall broader impact of action), (ii) purpose (immediate outcome/s), (iii) outputs (specific deliverable results to achieve objectives), and (iv) activities (key processes to be undertaken to produce the expected results). On the other side, the log frames provided; (i) verifiable indicators relating to the overall goal, purpose, outputs, key activities and inputs, (ii) means of verification (sources of information), and (iii) important assumptions (external conditions) [85]. Involvement of multiple stakeholders was required to assess the likelihood of implementing feasible actions. Therefore, stakeholders’ resolutions determined the applicability and acceptability of such a framework at sub-national levels of PC-NTD endemic settings in Kenya.

### Strengths and limitations

A key strength of developing the framework lies within the third phase involving a Delphi survey, which identified the implementable actions considering health personnel perspectives. Therefore, the framework components were informed by feasible recommendations to improve surveillance and response to PC-NTDs at the sub-national levels. Validation of the framework involving relevant stakeholders was crucial and identified further opportunities for improving the framework components and assessing adoptability at the sub national level. However, to operationalise the framework, an in depth understanding of the decision making process at the sub-national level is vital to identify who makes decisions, what information is required to make decisions, and how and when decisions are to be made. The framework postulates improving existing surveillance systems considering both an integrated and targeted process to controlling PC-NTDs. However, it may only apply to regions co-endemic of at least two or more PC-NTDs since the framework components intend to guide the decision-making process considering an integrated disease control approach. Furthermore, the framework validation phase was undertaken amid the COVID-19 pandemic, which resulted in conducting the validation process while strictly adhering to the Kenya Ministry of Health COVID-19 guidelines. This limited the number of face-to-face sessions undertaken and restricted the number of participants present within a given session. However, further resolutions from stakeholders were obtained through adopting alternative virtual mechanisms.

## Conclusion

Framework development was based on inclusion of factors that were initially assessed by key stakeholders regarding their importance and feasibility at the sub-national level. Further validation of the framework through a consultative process with concerned stakeholders aided in assessing its adoption, acceptability and utilisation for decision-making to improve PC-NTDs surveillance and response. However, acceptability of novel surveillance and response approaches are dictated by the socio-cultural and political dynamics that should be considered when instituting control efforts within a specific unique setting [16]. The ultimate goal of the framework is to contribute to efficient implementation of two core medical interventions of the five interventional packages outlined in the WHO work plans for NTDs elimination in Africa [8, 67]. The framework is inclined towards supporting implementation of interventions relating to PC for populations at risk of the infection and CF and IDM, which are critical to achieving PC-NTDs elimination. Attaining disease elimination and eventual eradication requires effective routine surveillance systems for early detection of possible instances of disease re-emergence both nationally and sub-nationally. However, health system strengthening frameworks in resource-constrained settings have predominantly focused on national levels with minimal focus on sub-national levels [86]. Therefore, the validated framework intends to guide decisions that will direct efforts towards assessing surveillance system performance at the sub-national level in view of PC-NTDs and inform implementation of relevant policies and control programmes.

## Data Availability

The datasets generated and/or analysed in the manuscript are not publicly available due to the need to keep the identities of participants confidential as they granted consent to be enrolled in the overall study on the basis of remaining anonymous, but are available from the corresponding author on reasonable request.

## Acknowledgements

The authors acknowledge the Kenya Ministry of Health and the County level authorities for granting permission to undertake the prior surveys that informed framework development. We also extend our sincere gratitude to all the healthcare personnel who participated in the surveys and framework validation process.

## Author Contributions

AKSN: conceived the research idea and designed the study, conducted data collection, data analysis, developed the draft framework and drafted the manuscript.

CMM and KV: provided supervision, intellectual input and guidance for developing the framework and drafting the manuscript.

All the authors read and approved the final version of the manuscript before submission.

